# Self-blaming emotions in major depression: a randomised pilot trial comparing fMRI neurofeedback training with self-guided psychological strategies (NeuroMooD)

**DOI:** 10.1101/19004309

**Authors:** Tanja Jaeckle, Steven C.R. Williams, Gareth J. Barker, Rodrigo Basilio, Ewan Carr, Kimberley Goldsmith, Alessandro Colasanti, Vincent Giampietro, Anthony Cleare, Allan H. Young, Jorge Moll, Roland Zahn

**Affiliations:** Department of Psychological Medicine, Centre for Affective Disorders, Institute of Psychiatry, Psychology & Neuroscience, King’s College London, United Kingdom; Department of Neuroimaging, Institute of Psychiatry, Psychology & Neuroscience, King’s College London, United Kingdom; Cognitive and Behavioral Neuroscience Unit and Neuroinformatics Workgroup, D’Or Institute for Research and Education (IDOR) – Rio de Janeiro, Brazil; Department of Biostatistics, Institute of Psychiatry, Psychology & Neuroscience, King’s College London, United Kingdom

**Keywords:** real-time fMRI, fMRI neurofeedback, clinical trial, guilt, major depressive disorder, anger, subgenual cingulate cortex, subgenual cortex, BA 25, anterior temporal lobe, social cognition, psychotherapy, cognitive behavioural therapy

## Abstract

**Background:** Overgeneralised self-blame and worthlessness are key symptoms of major depressive disorder (MDD) and were previously associated with self-blame-selective changes in connectivity between right superior anterior temporal lobe (rSATL) and subgenual frontal areas. In a previous study, remitted MDD patients successfully modulated guilt-selective rSATL-subgenual cingulate connectivity using real-time functional magnetic resonance imaging (rtfMRI) neurofeedback training, thereby increasing their self-esteem. The feasibility and potential of using this approach in symptomatic MDD were unknown.

**Methods:** This single-blind pre-registered randomised controlled pilot trial tested the clinical potential of a novel self-guided psychological intervention with and without additional rSATL-posterior subgenual cortex (SC) rtfMRI neurofeedback, targeting self-blaming emotions in insufficiently recovered people with MDD and early treatment-resistance (n=43, n=35 completers). Following a diagnostic baseline assessment, patients completed three self-guided sessions to rebalance self-blaming biases and a post-treatment assessment. The fMRI neurofeedback software FRIEND was used to measure rSATL-posterior SC connectivity, while the BDI-II was administered to assess depressive symptom severity as a primary outcome measure.

**Results:** Both interventions were demonstrated to be safe and beneficial, resulting in a mean reduction of MDD symptom severity by 46% and response rates of more than 55%, with no group difference. Secondary analyses, however, revealed a differential response on our primary outcome measure between MDD patients with and without DSM-5 defined anxious distress. Stratifying by anxious distress features was investigated, because this was found to be the most common subtype in our sample. MDD patients without anxious distress showed a higher response to rtfMRI neurofeedback training compared to the psychological intervention, with the opposite pattern found in anxious MDD. We explored potentially confounding clinical differences between subgroups and found that anxious MDD patients were much more likely to experience anger towards others as measured on our psychopathological interview which might play a role in their poorer response to neurofeedback. In keeping with the hypothesis that self-worth plays a key role in MDD, improvement on our primary outcome measure was correlated with increases in self-esteem after the intervention and this correlated with the frequency with which participants employed the strategies to tackle self-blame outside of the treatment sessions.

**Conclusions:** These findings suggest that self-blame-selective rtfMRI neurofeedback training may be superior over a solely psychological intervention in non-anxious MDD, although further confirmatory studies are needed. The self-guided psychological intervention showed a surprisingly high clinical potential in the anxious MDD group which needs further confirmation compared versus treatment-as-usual. Future studies need to investigate whether self-blame-selective rSATL-SC connectivity changes are irrelevant in anxious MDD, which could explain their response being better to the psychological intervention without interfering neurofeedback.

**https://doi.org/10.1186/ISRCTN10526888**

## Introduction

Recent findings confirm cognitive models (Abramson, Seligman & Teasdale, 1978) highlighting the importance of overgeneralised self-blaming emotions for major depressive disorder (MDD) vulnerability (Green, Moll, Deakin, Hulleman, & Zahn, 2013; Zahn et al., 2015a; Zahn et al., 2015b). Using fMRI, abnormal functional connectivity between the right superior anterior temporal lobe (rSATL) and the anterior subgenual cingulate cortex was found to be associated with overgeneralised self-blaming emotions in remitted MDD (Green, Lambon Ralph, Moll, Deakin & Zahn, 2012). Increased functional connectivity in remitted MDD between the rSATL and posterior subgenual cortex (SC) predicted risk of future major depressive episodes (MDEs) over the subsequent year (Lythe et al., 2015). Whereas Sato et al. (2013) provided the technical proof-of-concept that changes in selective functional connectivity can be detected and fed back to healthy control (HC) participants during fMRI scanning, a recently completed double-blind, randomised clinical trial confirmed that fMRI neurofeedback can successfully train remitted MDD patients in rebalancing abnormal brain connectivity patterns (Zahn et al., 2018).

Real-time fMRI neurofeedback is a training method that provides the individual with near real-time information about changes in neural activity to facilitate self-regulation of brain function, cognition and behaviour (Stoeckel et al., 2014; Thibault, Lifshitz, & Raz, 2016). It is a recent and less widely used experimental approach, yet using this technique, it has been demonstrated that individuals learn quickly to gain voluntary control over the activation and connectivity of specific brain regions (Sulzer et al., 2013; Weiskopf, 2012). Only a few studies to date have administered rtfMRI neurofeedback in patients with depression (Young et al., 2017a; Young et al., 2018a; Young et al., 2018b; Yuan et al., 2014; Zotev, Phillips, Yuan, Misaki, & Bodurka, 2014; Zotev et al., 2016), and even fewer studies investigated the use of rtfMRI neurofeedback as a therapeutic intervention strategy in MDD (Linden et al., 2012; Mehler et al., 2018; Young et al., 2017b; Young et al., 2014). Linden et al.’s (2012) pioneering study applied rtfMRI neurofeedback training targeted at increasing activation in brain areas involved in the processing of positive emotions, i.e. the ventrolateral prefrontal cortex and insula, whereas Young et al. (2014) used neurofeedback training to enhance amygdala response during the recall of positive autobiographical memories. Both studies assessed whether the rtfMRI neurofeedback interventions would have a significant effect on symptom severity in MDD as assessed with the 17–item Hamilton Rating Scale for Depression (Linden et al., 2012) and Profile of Mood States (POMS) depression ratings (Young et al., 2014). Although both studies delivered promising results and found significant reductions in symptom severity, it is noteworthy that they lacked randomisation and employed only small sample sizes (n=8 vs n=8 controls (Linden et al., 2012) and n=14 vs n=7 controls (Young et al., 2014)).

A more recently published study by Young et al. (2017a) was the first randomised rtfMRI neurofeedback trial in MDD, investigating medication-free individuals allocated to moderately sized groups (n=19 vs n=17 controls). Similarly, to the authors’ previous research approach (Young et al., 2014), rtfMRI neurofeedback was used to increase the individual’s amygdala hemodynamic response to positive autobiographical memories. Symptom reduction was assessed using the Montgomery-Åsberg Depression Rating Scale (MADRS). Young et al. (2017a) observed a significant symptom reduction in patients allocated to the active neurofeedback group compared to a minimal response in the control neurofeedback group. Contrary to Young et al.’s results (2017a), another randomised controlled rtfMRI neurofeedback trial conducted by Mehler et al. (2018) did not find differences between the active and control rtfMRI neurofeedback MDD group. Interestingly, both rtfMRI neurofeedback groups successfully upregulated targeted brain areas and reduced their depression symptoms by more than 40%. Further, patients’ symptoms remained stable at a follow-up assessment six weeks after trial completion (Mehler et al., 2018).

It is noteworthy that most previous clinical rtfMRI neurofeedback studies focussed on investigating remission from depressed states rather than early treatment resistance or recurrence risk. Apart from recent work conducted by Mehler et al. (2018), to our knowledge, no rtfMRI neurofeedback intervention has been developed to this date which aims to reduce symptoms in MDD patients who have only insufficiently responded to standard treatment, a clinical predictor of recurrence risk in MDD. It has not been explored yet if the clinical benefits of rtfMRI neurofeedback as a therapeutic tool are more profound in those patients who only insufficiently respond to standard treatment, which is the reason why the NeuroMooD trial was conducted in early treatment-resistant MDD patients. Given that self-blame-selective hyper-connectivity between the rSATL and the posterior SC (Brodmann Area [BA] 25) predicted recurrence risk in MDD (Lythe et al., 2015), this was used as our target in the rtfMRI neurofeedback intervention in the current trial.

### NeuroMooD trial: aims and objectives

Building on previous research findings as outlined above and grounded in the need for novel intervention strategies in the treatment of early treatment-resistant MDD, this research investigated and compared two novel approaches.

Specifically, this clinical trial examined the clinical benefits of a novel rtfMRI neurofeedback protocol in current and insufficiently remitted MDD, aiming at self-blame-selective neural connectivity abnormalities between the rSATL and the posterior SC. The therapeutic effects of this rtfMRI neurofeedback intervention were compared to the proposed benefits of a newly designed, self-guided psychological intervention. Clinically, both interventions aimed at alleviating symptoms of depression, and effectively reducing self-blaming emotions, in addition to improving the sense of self-worth. Moreover, specific to the rtfMRI neurofeedback condition, the aim was to determine whether rtfMRI neurofeedback training effects on excessive self-blame and depressive symptoms were associated with the normalisation (i.e. decrease) of self-blame-selective hyper-connectivity between the rSATL and the posterior SC (BA 25).

### Hypotheses

<u>The following hypotheses were investigated:</u>

#### Pre-registered Main Hypothesis 1

Patients undergoing rtfMRI neurofeedback training will show reduced depressive symptoms, decreased self-blame and increased self-worth when compared with the psychological intervention group.

#### Specific Secondary Hypothesis 2

Patients undergoing rtfMRI neurofeedback training will show decreased self-blame-selective hyper-connectivity between the rSATL and the posterior SC post-treatment compared to pre-treatment (one of our pre-registered secondary outcome measures).

#### Specific Secondary Hypothesis 3

Decreased self-blame-selective hyper-connectivity between the rSATL and the posterior SC region is associated with a reduction in depressive symptoms in MDD.

## Methods

This clinical proof-of-concept trial received ethical approval from the NHS Health Research Authority, NRES Committee London – Camberwell St Giles (REC reference: 15/LO/0577) and was pre-registered on the ISRCTN registration database (ISRCTN10526888). The single research site was the Institute of Psychiatry, Psychology & Neuroscience, King’s College London.

Researchers involved in the conduction of this clinical trial affirm that study procedures complied with the ethical principles, standards and national and institutional guidelines for clinical trials and research involving human subjects and with the Helsinki Declaration of 1975, as revised in 2008.

### Trial design

A single-blind, randomised controlled trial design was used, and participants allocated to two distinct treatment arms, each comprising three intervention visits (visits 2, 3 & 4). Regardless of the intervention group, treatment sessions were scheduled 7-13 days apart, depending on the participants’ availability.

Feasibility and effectiveness of both interventional approaches were compared by measuring the change in clinical outcomes between pre-treatment (visit 1) and post-treatment assessments (visit 5).

One intervention condition implemented three sessions of a self-guided psychological intervention that consisted of cognitive reappraisal techniques, modified from cognitive therapy (Beck, Rush, Shaw &Emery, 1979) and related approaches. Assigned to the second intervention condition, the rtfMRI treatment group, participants were asked to apply the same self-guided psychological strategies during three sessions of rtfMRI neurofeedback training, targeting rSATL-posterior SC correlation.

Initially the NeuroMooD study was designed to compare three treatment arms, investigating the treatment effects of rtfMRI neurofeedback with an active, cathodal and a sham transcranial direct current stimulation (tDCS) intervention. Specifically, the original single-blind, randomised controlled design compared three sessions of the rtfMRI neurofeedback treatment as described above with three sessions of right superior temporal lobe cathodal tDCS plus self-guided psychological intervention and three sessions of sham right superior temporal lobe tDCS plus self-guided psychological intervention. Due to funding reasons, the original trial design had to be modified, and the data of 6 randomised participants, that had already been collected, were discarded.

### Randomisation method

The randomisation of trial participants was performed by an automatised online system, set up by the Clinical Trials Unit, King’s College London. The randomisation process implied a stratified block design with randomly varying block sizes, deploying two stratification factors: gender (female/male) and baseline scores of the primary outcome measure, the Beck Depression Inventory-II (BDI-II; Beck, Steer & Brown, 1996). Baseline scores classified participants based on designated BDI-II categories of symptom severity as follows: BDI-II scores below 14 points indicating minimal depression, BDI-II scores between 14 and 28 points comprising mild and moderate depression and BDI-II scores of 28 points or higher, implying severe depressive symptoms. Participants were informed about their allocated treatment group upon completion of the baseline clinical and neuropsychological testing on their pre-treatment assessment (visit 1).

### Recruitment and reimbursement of participants

The recruitment/randomisation phase consisted of a total of 15 months, from September 2016 to December 2017. Trial adverts were posted primarily online, further recruitment strategies entailed the dissemination of study adverts via university and institutional recruitment circulars, as well as presenting to self-help groups at scheduled member meetings.

Participants received compensation for the time taken to participate in the study in the form of high street gift vouchers or shopping vouchers. Reimbursement was appointed on a pro-rata basis on the final day of participation: vouchers worth £10 for the pre-trial assessment session (visit 1), vouchers worth £20 per treatment session (3 x £20 = £60 for visit 2, visit 3 and visit 4). Additional vouchers worth £30 for the final follow-up session (visit 5).

### Inclusion criteria

Recruitment for this clinical trial was targeted at patients suffering from recurrent MDD according to diagnostic criteria of the Diagnostic and Statistical Manual of Mental Disorders (DSM-5; American Psychiatric Association [APA], 2013), with a minimum of one past MDE of at least a two months duration. At baseline assessment (visit 1), patients either currently experienced an MDE or had insufficiently recovered, presenting with significantly impairing or bothering symptoms, despite not fulfilling MDE criteria anymore. It was required that remaining symptoms would be significant in severity, classified as a psychiatric status rating of three (i.e. significant symptoms) or four (i.e. major symptoms) on the Longitudinal Interval Follow-up Interview (LIFE; Keller et al., 1987) over the past two weeks prior baseline assessment and randomisation. Further, MDD patients needed to be stable in symptoms for at least six weeks before randomisation to minimise the risk of including spontaneous remitters.

Importantly, MDD patients were required to have shown an only insufficient response to at least one psychological intervention (e.g. cognitive behavioural therapy) or antidepressant medication before their enrolment in the study or were not amenable to these standard forms of treatment. MDD patients could only be included if they were not currently undergoing psychotherapeutic treatment. Antidepressant medication was no exclusion criterion, but patients needed to be on a stable dose for at least six weeks without improvement before their participation and were asked to remain on this dose throughout the study. Lastly, participants needed to be aged 18 years or older, right-handed (to ensure homogenous responses to the right hemispheric treatment target), and be proficient in English, so that reliable responses on newly developed secondary outcome measures could be collected.

### Exclusion criteria

To ensure safety and minimise potential health risks, participants needed to be excluded if they presented with greater than a low risk of suicidality, violence or current self-harming behaviour. Additionally, participants presenting with a current MDE lasting more than 12 months were excluded.

Additional exclusion criteria were defined as follows:

i. Standard MRI contraindications, i.e. non-removable ferromagnetic devices or implants due to the possible dangerous effects of the MRI magnet upon metal objects in the body
ii. History of manic or hypomanic episodes, of schizophreniform symptoms or schizophrenia, or substance abuse
iii. History of neurological disorders such as seizures, loss of consciousness following brain injury or medical disorders affecting brain function, blood flow or metabolism
iv. History of learning disabilities, major medical, developmental or relevant other axis-I disorders
v. Prior specialist diagnosis of attention deficit hyperactivity disorder (ADHD), antisocial or borderline personality disorder
vi. Significant impairment of psychosocial functioning before the last MDE indicating the possibility of a comorbid personality disorder
vii. Current intake of benzodiazepines, GABAergic or benzodiazepine receptor agonists
viii. Current recreational drug use
ix. Past violence or current aggressive impulses
x. Impairments of vision or hearing which cannot be corrected during the treatment sessions
xi. Pregnancy

### Assessment and evaluation of participants: eligibility assessment

The participant selection process commenced with a telephone-based screening of volunteers for inclusion and exclusion criteria after they gave oral informed consent, they then provided written informed consent after passing the pre-screening stage. During the recruitment phase, a total of 311 volunteers interested in participating in the study were screened over the phone, and 71 volunteers attended the initial baseline assessment (visit 1). Following diagnostic and clinical evaluation, ultimately, N=43 participants were randomised into the study, of which n=35 participants completed this clinical trial.

### Assessment and evaluation of participants: clinical assessment

The diagnostic, clinical and cognitive assessment comprised standardised, validated measures that have been used extensively in psychiatric research.

### Summary of standard clinical and cognitive instruments

i. Structured Clinical Interview (SCID) for DSM-5 (First, 2015)
ii. AMDP Psychopathology Interview questions on depression (Faehndrich & Stieglitz, 1997; Zahn et al., 2015b)
iii. Longitudinal Interval Follow-Up Evaluation (LIFE; Keller et al., 1987)
iv. Clinical Global Impression (CGI) Scale (Busner & Targum, 2007)
v. Beck Depressive Inventory (BDI-II; Beck et al., 1996)
vi. Montgomery-Åsberg Depression Rating Scale (MADRS; Montgomery & Åsberg, 1979)
vii. Quick Inventory of Depressive Symptomatology (QUIDS-SR16; Rush et al., 2003)
viii. Hypomania Checklist-16 (Forty et al., 2010)
ix. Rosenberg Self-Esteem Scale (Rosenberg, 1965)
x. Profile of Mood States (POMS) Scale (McNair, Lorr & Dropplemen, 1971)
xi. MINI International Neuropsychiatric Interview (module on suicidality only; Sheehan et al., 1998)
xii. Psychiatric Family History Screen (Weissman et al., 2000)
xiii. Life Events Questionnaire (Brugha & Conroy, 1985)
xiv. Childhood Trauma Questionnaire (CTQ; Bernstein & Fink, 1998)
xv. Altman Self-Rating Mania Scale (Altman, Hedeker, Peterson & Davis, 1997)
xvi. Addenbrooke’s Cognitive Examination (ACE-III; Hsieh, Schubert, Hoon, Mioshi & Hodges, 2013) in patients >50 years only

In addition to the abovementioned scales and measures, clinical evaluation of participants further entailed a non-structured clinical interview, as well as the documentation of the patient’s medical history and in females the day in their menstrual cycle. Furthermore, age at onset, episode duration(s), and total illness duration was recorded, along with details about the course of illness, i.e. number of episodes and medication history.

### Additional experimental neuropsychological testing

Supplementary to clinical and cognitive assessments, participants were asked to provide ratings of autobiographical memories associated with feelings of self-blame and other-blame. Moreover, additional experimental tasks developed by our research group were administered, designed to explore neurocognitive aspects of implicit self-contempt biases and self- and other-blaming emotions:

i. A modified short version of the value-related moral sentiment task (VMST; Zahn et al., 2015a): this computerised task investigates emotions related to self-blame (i.e. guilt, shame, self-contempt, self-disgust, self-directed anger) versus blaming others (indignation, anger, contempt or disgust towards others). Preceded by the description of hypothetical scenarios of social behaviours of the participants themselves and their best friends, participants are instructed to select the emotion they are most likely to experience. We added items related to action tendencies (Roseman, Wiest & Swartz, 1994), previously validated in an unpublished study. The following action tendencies were measured: creating distance from self, hiding, apologising, creating distance from friend, verbally or physically attacking/punishing friend or no action/other action. For this trial report we only focus on the pre-registered secondary outcome measure of agency-incongruent self-blaming emotions, so the percentage of other-agency trials where shame, guilt, or self-disgust/contempt were experienced. This was chosen because unpublished analyses of previous data showing a correlation of rsATL-posterior SC connectivity for self-blame vs. other-blame and agency-incongruent self-blaming emotions on the full version of the VMST (Lythe et al., 2015).
ii. Brief Implicit Association Test (BIAT; Sriram & Greenwald, 2009): this computerised task using Inquisit Software (www.millisecond.com) was developed by our research group by RZ and Dr Karen Lythe in collaboration with Profs Rüsch and Bodenhausen. It is an indirect measure of self-contempt bias, evaluating the association of contempt or disgust with oneself relative to others. The task design is based on similar tests that have been validated to measure implicit self-esteem (Greenwald & Farnham, 2000).
iii. A modified version of the social knowledge differentiation task (Green et al., 2013): this computerised, neuropsychological test examines the participant’s ability to access differentiated social conceptual knowledge when instructed to appraise hypothetical scenarios of social behaviour of different contexts of agency (self-agency vs other-agency). The task was modified by restricting the original task to 30 items, focussing on negatively valenced scenarios only. This was not a pre-registered secondary outcome measure and the results will be reported in a separate paper.
iv. Social agency inference task (SAIT): Specifically developed for this research project, this computerised task assesses whether changes in the perception of social agency underpin self-blaming biases in MDD. This task was not a pre-registered secondary outcome measure and will be reported in a separate paper.

### Pre-registered outcome measures

Outcome measures comprised self-rated and observer-rated scales and assessments along with fMRI connectivity analyses as specified below. Observer-rated outcomes were assessed by a senior psychiatrist (R.Z. or A.C. in his absence) who was blinded to the treatment group allocation of participants throughout the trial.

### Primary outcome measure

The primary outcome measure was defined as the reduction of depressive symptoms between pre-treatment visit 1 and post-treatment visit 5 (7-13 days after final treatment session) as assessed with the BDI-II.

### Secondary outcome measures

In addition to the primary outcome measure, the following secondary outcome measures had been pre-registered before the begin of the trial:

i. Reduction of depressive symptoms between pre-treatment visit 1 and post-treatment visit 5 (7-13 days after final treatment session) as assessed with MADRS
ii. Reduction of self-rated depressive symptoms between pre-treatment visit 1 and post-treatment visit 5 (7-13 days after final treatment session) as assessed with QUIDS-SR16
iii. Reduction of self-rated depressive symptoms between pre-treatment visit 1 and post-treatment visit 5 (7-13 days after final treatment session) as assessed with POMS depression-dejection subscale
iv. Increase in self-worth between pre-treatment visit 1 and post-treatment visit 5 (7-13 days after final treatment session) as assessed with Rosenberg Self-Esteem Scale
v. In the rtfMRI neurofeedback group: decrease in post vs pre-training rSATL–posterior SC correlation for self-blame relative to blaming others between the first and last treatment session (i.e. at the start of visit 2 and at the end of visit 4), using fMRI as measured by regression coefficients for the time series, as extracted by the software FRIEND (Functional Real-time Interactive Endogeneous Neuromodulation and Decoding; Basilio et al., 2015; Sato et al., 2013)
vi. Reduction in implicit self-blaming bias between pre-treatment visit 1 and post-treatment visit 5 (7-13 days after final treatment session) as assessed with BIAT (subcategories contempt–anger and contempt–anxiety)
vii. Reduction in agency-incongruent self-blame between pre-treatment visit 1 and post-treatment visit 5 (7-13 days after final treatment session) as assessed with the short version of the VMST
viii. Self- and observer-rated clinical global impression at post-treatment visit 5 (7-13 days after final treatment session) as assessed with the CGI-Scale
ix. Withdrawal rates throughout the trial and separately for the period after the first treatment session until post-treatment visit 5 (7-13 days after final treatment session)
x. Adverse events throughout the trial and separately for the period after the first treatment session until post-treatment visit 5 (7-13 days after final treatment session)
xi. Reduction in self-rated self-blame as assessed with the mean of self-blame ratings of two guilt-specific autobiographical events obtained prior the first (visit 2) and after final treatment session (visit 4)
xii. Reduction in observer-rated self-blame as assessed with the Moral Emotion Addendum to the AMDP as the sum of all self-blaming emotion scores at baseline (visit 1) and post-treatment at visit 5 (7-13 days after final treatment session)

### Intervention procedures

Both interventions, the psychological intervention as well as the rtfMRI neurofeedback training, consisted of three individual treatment sessions, scheduled 7-13 days apart, and involved equivalent preparation processes prior to the first treatment session (see participant timing in Supplementary Table 1).

Before the first intervention session, participants were asked to provide two cue words, prompting them to remember two autobiographical events that would cause them to experience strong feelings of self-blame and guilt. Also, participants provided two cue words reminding them of life events where they experienced substantial feelings of indignation or anger towards other people while feeling low levels of self-blame.

Before and after each treatment session, participants rated the intensity of evoked self-blame and indignation feelings on a Likert-type scale from 0 to 10. Moreover, they rated (from 0 to 10) how successful they felt in the emotional training during the intervention and estimated the percentage of time (0-100%) that they were able to focus during the session.

Concluding each treatment session, the participant’s suicide risk was assessed using the MINI International Neuropsychiatric Interview suicidality module, focussed on the time period since the previous study appointment. In addition, the severity of depressive symptoms was monitored and assessed with the BDI-II. Participants were excluded if they showed a suicide risk greater than low on the MINI and as judged by R.Z., or if their depressive symptoms had worsened, as reflected in an increase of 10 points or more on the BDI-II compared to the baseline score prior randomisation. In such an instance, the protocol requested to un-blind the leading senior psychiatrist of the NeuroMooD trial (R.Z.), who discussed treatment recommendations with the patient if requested. By doing so, participants were assisted in accessing standard treatment options swiftly.

### Psychological intervention

In the psychological intervention group, the cue words provided by participants of this treatment arm were programmed into a timed presentation format and played back to them during each treatment session. To help participants manage their feelings of self-blame constructively, they were instructed to use specific, self-guided psychological strategies. Participants were suggested to use the following strategies to help them manage their feelings, yet they could also develop their own strategies:

i. Think about why you might not have been in control over the outcome of the event.
ii. Think about why you might not be responsible for the outcome of the event.
iii. Think about why the consequences for others might not be so bad.
iv. Think about making up for things or apologising.
v. Think about the other person forgiving you.
vi. Think about forgiving yourself.

These strategies were based on (1) attribution theory which highlights the importance of locus of control for self-blame (Abramson et al., 1978), on (2) omnipotent responsibility associated with depressogenic forms of guilt (O’Connor, Berry, Weiss, & Gilbert, 2002), on (3) neurocognitive models of self-blame, implicating representations of future consequences as important to guilt-proneness (Zahn R, de Oliveira-Souza & Moll, 2013), on (4) the associations of reparative action tendencies with adaptive forms of guilt (Tangney, Stuewig, & Mashek, 2007), as well as on (5) the focus on forgiveness and self-kindness as thematised in compassion-focused therapy (CFT; Gilbert, 2009a, 2009b; Gilbert & Procter, 2006).

The intervention consisted of four parts. In the first and the fourth part, participants were asked to only think about the autobiographical events triggered by their cue words, without using any strategies to manage their feelings of self-blame. Before the second and third part of the intervention, participants were instructed to start using one or more self-guided strategies when seeing their guilt cue words to manage their feelings of self-blame constructively.

Participants were given the following instructions before the treatment session was started:

> ‘At the beginning and end of the session, you will have to think about the self-blame and anger events when shown your cue words. In between, you will be asked to keep thinking about the self-blame event while trying to use one or more strategies that best help you to cope with the self-blaming feeling. When numbers are presented on the screen, you will have to subtract seven from the number displayed.’

In the first and fourth part of the presentation, participants were shown their guilt, and their indignation cue words, parts two and three only contained the patient’s guilt cue words and no indignation provoking cue words. Parts 1 and 4 were 408 seconds in length, including emotional blocks (guilt and indignation) and subtraction blocks, plus a 30-second reminder of task instructions. Parts 2 and 3 consisted of a time sequence of 424 seconds each, containing guilt cue words and subtraction blocks, in addition to the display of instruction slides for 60 seconds. Consequently, the intervention part of each treatment session was completed after approximately 30 minutes.

The order of the displayed cue words and numbers was as follows:

Part 1 of the intervention: instruction to only think about the autobiographical events without using psychological strategies ➔ number ➔ guilt cue word 1 ➔ number ➔ indignation cue word 1 ➔ number ➔ guilt cue word 1 ➔ number ➔ indignation cue word 1 ➔ number ➔ guilt cue word 2 ➔ number ➔ indignation cue word 2 ➔ number ➔ guilt cue word 2 ➔ number ➔ indignation cue word 2.

Part 2 of the intervention: instruction to keep thinking about the events, while trying to use psychological strategies to cope with self-blaming feeling ➔ number ➔ guilt cue word 1 ➔ number ➔ guilt cue word 1 ➔ number ➔ guilt cue word 2 ➔ number ➔ guilt cue word 2.

Part 3 was equal to part 2 of the intervention. Part 4 of the intervention was identical to Part 1.

The mental subtraction blocks served as a distraction from the emotional load of the participants’ thought processes and to separate each emotional block. Self-blame cue words were presented in blue colour on a black background; indignation cue words appeared in red on black background and numbers were presented in yellow.

In both treatment groups, participants were instructed to implement the psychological strategies in their everyday lives and to use them in-between treatment visits whenever feelings of self-blame would arise. The frequency of use was recorded at the next treatment visit. Participants were instructed to continue using the strategies until the final assessment visit (visit 5).

### Real-time fMRI neurofeedback intervention

The rtfMRI neurofeedback intervention aimed at targeting hyper-connected brain correlation patterns between the rSATL seed and the posterior SC region of interest (ROI) for self-blame vs. blaming others, identified as a signature of vulnerability to MDD (Lythe et al., 2015).

Analogous to the psychological intervention group, and before the first neurofeedback training session, participants were asked to decide on specific autobiographical memories that would evoke strong feelings of self-blame and other-blame when prompted by previously defined cue words. The self-blame-evoking scenarios had to involve the participant as the main agent of the scenario. The other-blaming scenarios had to involve another person acting. To evaluate whether a change occurred in the attribution of blame, ratings on these events were obtained before and after each scanning session. Instructions were given through the MRI intercom, participants, however, responded with a button box to prevent extensive head movement while being in the scanner.

Analogous to the psychological intervention group, each of the three rtfMRI neurofeedback sessions contained a paradigm of four runs, whereby the following procedure applied:

The first and fourth run (204 volumes each; 408 seconds duration) were identical and served to determine pre- and post-neurofeedback effects. They constituted of rtfMRI data acquisition runs, consisting of four self-blame (guilt) blocks (15 volumes each) and four other-blame (indignation) blocks (15 volumes each), interspersed with eight mental subtraction condition blocks (10 volumes each). As mentioned earlier, during the subtraction blocks, participants were asked to mentally subtract seven from a 3-digit number (e.g. 101, 102).

While run 1 measured the correlation coefficient of self-blaming emotions relative to other-blaming emotions (indignation), training effects on such correlations were assessed in run 4.

During the neurofeedback training runs (run 2 and 3), an upward and downward moving thermometer scale was displayed to provide visual feedback on how successful participants were in modifying their brain correlation patterns between the rSATL and the posterior SC region ROI. The thermometer scale appeared in the form of a colour bar that could reach different levels. Participants were instructed to think about the particular autobiographical scenario triggered by the display of the previously agreed cue word and to try and bring up the level to the top of the thermometer scale by using choosing a psychological strategy from the list they had been provided with before the scanning session.

Runs 2 and 3 (212 volumes each; 424 seconds duration) were identical and consisted of four guilt blocks (42 volumes per block), interspersed with four mental subtraction condition blocks (10 volumes each).

Similar to the psychological intervention group, mental subtraction blocks were used to divert participants from the emotionally charged autobiographical memories, in addition to minimising resting-state activity in the posterior SC region (Bado et al., 2014).

### Real-time fMRI neurofeedback method

The rtfMRI neurofeedback software FRIEND (Basilio et al., 2015; Sato et al., 2013) was used in the file version 1.0.0.257 (Supplementary Figures 1&2). FRIEND has previously been validated for correlation feedback in patients with MDD (Zahn et al., 2018).

A detailed description of methodological specifications of FRIEND as a rtfMRI neurofeedback tool is provided by Sato et al. (2013) and Zahn et al. (2018). In the NeuroMooD trial, FRIEND provided ROI-based rtfMRI neurofeedback alongside executing fundamental pre-processing steps of fMRI data in real-time. Facilitated by native FSL codes, FRIEND performed motion correction using MCFLIRT, spatial smoothing with Gaussian Kernel (FWHM = 6mm) and GLM calculation (Zahn et al., 2018).

Signal-level normalisation was performed by subtracting the mean value of the voxels signals within the ROI over the entire preceding subtraction condition block from the current echo-planar images belonging to the guilt or indignation condition block, which minimises local signal trends (Zahn et al., 2018).

The rSATL ROI (consisting of the same region used as a seed region in our previous studies (Green et al., 2012)) and posterior SC ROI (consisting of the BA 25 cluster whose self-blame-selective hyper-connectivity was associated with recurrence risk (Lythe et al., 2015)) were pre-defined, warped from MNI space into subject space and ultimately back-transformed into native space, using inverse transformation algorithms of FSL FLIRT (affine, 12 parameters). During run 1, 50% of the most activated voxels were selected in the native space ROI, contrasting the activation between guilt vs subtraction in the rSATL ROI, while contrasting guilt vs indignation in the posterior SC ROI. These voxels were used to extract the average signal for the subsequent rtfMRI neurofeedback training. The first five volumes of each emotional block were discarded due to high correlations guided by a decrease in time series after subtraction conditions (Zahn et al., 2018).

Thermometer levels, as displayed in the neurofeedback training runs, were calculated from the participant’s correlation patterns with a delay of six seconds. Once the first ten time points had been acquired to compute a correlation coefficient, the thermometer was updated every two seconds as soon as a new time point had been collected (i.e. each acquired volume).

FRIEND used a moving target correlation algorithm over a sliding time window of the last ten volumes, updated every two seconds, hence for each acquired volume. The minimum of the thermometer display was calculated based on the minimum value of the last 10 Pearson correlation coefficients, whereas the maximum of the thermometer was calculated based on the maximum value of the last ten correlations.

### Image acquisition

Image acquisition was carried out on an MR750 3.0T MR system (General Electric), using a hyperbolic secant (HS) excitation pulse, optimised for orbitofrontal and inferior temporal regions, minimising signal dropout (Wastling & Barker, 2015). A 32-channel head coil was chosen to support an optimal signal-to-noise ratio. Functional image acquisition was obtained in the AC-PC plane, top to bottom, using a T2*-weighted echo-planar imagining EPI (BOLD) sequence (TR = 2000 ms, TE = 30 ms, matrix = 64×64, FOV = 211 mm, flip angle = 73°, voxel size = 3×3×3 mm; slice thickness =3 mm, slice gap = 0.3 mm, 36 slices). Auto shimming was applied before starting each experimental run, acquiring four additional volumes which were automatically discarded, accounting of T1 equilibration effects. High-resolution anatomical images were acquired with a magnetisation–prepared rapid gradient echo (MP-RAGE) sequence (TR = 7.3 sec, TE = 3.0 sec, matrix = 256 x 256, FOV = 270 mm, slice thickness = 1.2 mm, 196 slices).

Clinical images were acquired on the first day of treatment (visit 2) using an FRFSE (2 mm thickness, 72 slices) and FLAIR sequence (4 mm thickness, 36 slices) and checked for anatomical brain abnormalities after the treatment session by a radiologist at the Centre for Neuroimaging Sciences, King’s College London, independent of additional, internal checks completed by the NeuroMooD study team.

While being in the MRI scanner, the participant’s head motion was restricted using padding and heart rate measurements recorded via a finger pulse sensor. A mirror fitted to the head coil allowed MDD patients to view visual stimuli presented during image acquisition, i.e. autobiographical cue words and the visual feedback thermometer, as stimuli were projected to a screen located behind the participant’s head. Verbal instructions were communicated via the MRI intercom, participants, however, were instructed to respond using a button box placed in their hands to avoid incidental head movement.

### Statistical power and offline analyses

Statistical power was calculated using G*POWER software and required a sample size of n=34 participants to achieve 85% power at p=.05, 2-sided (t-test). This calculation was based on a conservatively estimated effect size (d=1.06) lower than the effect size (d=1.5) reported in a previous rtfMRI neurofeedback study in MDD (Linden et al., 2012). The enrolment target consisted of n=45 MDD patients, including a 20% drop-out rate of 9 participants. Hence, this trial aimed to conclude with n=36 MDD study completers, above n=34 as needed per power calculation.

Given that this clinical trial was conducted as a pilot study to test feasibility, the performed power calculation is of limited value as it is based on effect sizes which may have been inflated due to small sample sizes. To determine precise effect sizes, a feasibility study is needed which should include at least 70 participants (i.e. 35 participants per group) when estimating the pooled standard deviation for continuous outcomes in randomised controlled trials (Teare et al., 2014). Furthermore, based on guidelines posited by the National Institute for Health Research, feasibility studies should not intend to be based on standard power calculations (Teare et al., 2014). These strict guidelines, however, have not been adhered to by any neurofeedback study in MDD so far, due to the difficulty of obtaining funding for such large studies at an early stage of development.

Ultimately, n=43 participants were randomised into the study, whereby n=22 MDD patients were allocated to the fMRI neurofeedback group and n=21 to the psychological intervention group. A sum of 8 participants withdrew or was excluded during the duration of the trial, leading to a final of n=35 datasets for the analysis of the primary outcome measure (BDI-II). Further details about participant numbers and withdrawal rates are reported in Supplementary Figure 3.

Statistical analyses used IBM SPSS Statistics 24 (https://www.ibm.com/analytics/spss-statistics-software). Group level analyses of primary and secondary outcomes, comparing pre- and post-treatment effects (visit 1 vs visit 5), were obtained using the constrained longitudinal analysis model (cLDA; Coffman, Edelman & Woolson, 2016) after seeking statistical advice (K.G., E.C.), the alpha-level was set to p=.05, two-tailed.

As in our previous paper (Zahn et al., 2018) at the individual subject level, linear regression coefficients for the slope of z-transformed ATL signal time-course as the predictor of z-transformed SC signal time-course in each condition (self-blame, indignation/anger) in the pre- and post-training acquisition as the outcome variables were derived from a general linear model for each subject by modelling the interaction of z-transformed ATL signal time-course with two factors: condition (guilt, indignation) and time (pre-, post-training). The z-transformation was undertaken to obtain standardised regression coefficients.

Cohen’s d effect sizes were computed for each regression coefficient using the formula: 2 x t-value/square root of degrees of freedom (df; Rosenthal & Rosnow, 1991).

Where cLDA was inapplicable, intervention group comparisons were performed using non-parametric tests. A repeated measures ANOVA was chosen in the analysis of regression coefficients for z-transformed rSATL and posterior SC signals in the guilt and indignation conditions. Transforming the data into z-transformed values allowed for receiving standardised regression coefficients. Secondary data analyses of the anxious distress subtype of MDD were conducted using univariate GLM analysis. As analyses were either hypothesis-driven or exploratory (secondary outcome measures in the feasibility trial), p-value adjustments to correct for multiple comparisons were not carried out (Feise, 2002).

### Brief Implicit Association Test design and analyses

The Brief Implicit Association Test (BIAT) measures implicit associations between two categories compared with another pair using two blocks of 20 stimuli each (16 used for the analysis). Participants were asked to press a left or right button depending on their categorisation of a word appearing on the screen. The Contempt vs. Anxiety (non-focal category) BIAT presented words falling into self or other categories and contempt or anxiety categories. In Block 1: one response key had to be pressed if the word was either a synonym of contempt or of self, and the other key had to be pressed if it was either a synonym of anxiety or other. In Block 2, the assignment to the response keys was reversed with one key for contempt or other and one key for anxiety or self. The BIAT literature shows that people are faster during the block in which the pairing of categories to one key is congruent with their implicit association biases towards one pair of categories relative to the other pair of categories. The contempt vs. anger (non-focal category) BIAT used the same design, but anxiety was replaced by anger as a category. We computed D scores using the optimised scoring algorithm (Nosek, 2005, May 27) as response time means for Block 2 – means Block 1 / standard deviation across blocks. This means that participants with stronger biases towards associating contempt with self were expected to show faster responses in Block 1 and thus more positive BIAT D scores. Unlike other BIATs, we used self-agency and other-agency rather than just self and other (please see Supplementary Methods). Due to an undetected technical error in the set-up of the BIAT, we used “Participant” and “Friend” instead of the first names of participants and friends which may limit the validity of the results.

### NeuroMooD protocol violations

Minor violations of the NeuroMooD protocol occurred during the duration of this study due to difficulties in scheduling participants’ treatment and final assessment visits. Modified schedules had to be arranged for individual participants who were unable to attend study visits within the preferred interval of 7 to 13 days between appointments due to time constraints. Also, limited availability of fMRI scanning slots at the MRI facilities of the Centre for Neuroimaging Sciences, King’s College London, affected MDD patients allocated to the rtfMRI neurofeedback group, occasionally causing a delay in the scheduling of treatment visits. As this issue became apparent early in the study, treatment visits for the psychological treatment group were scheduled in intervals comparable to those of the rtfMRI neurofeedback group. Ultimately, no significant difference was found between treatment groups regarding the total number of days included in the study (t=1.21, df=33, p=.237, two-tailed), considering the period from randomisation (visit 1) until trial completion (visit 5). On average, it took rtfMRI neurofeedback participants 40 days (SD=9.18) to complete the NeuroMooD trial. Similarly, participants randomised to the psychological intervention group participated on average for 37 days (SD=7.37).

### Clinical characteristics of participants

## Results

### Pre-registered primary outcome measure

A significant improvement, irrespective of treatment group, was found on the pre-registered primary outcome measure, the BDI-II (Beck et al., 1996). A comparison of BDI-II score means pre- and post-treatment showed an overall reduction of 46.07%, which corresponds to a baseline assessment (N=43) of M=29.14 points (SD=8.66) and M=15.71, (SD=9.75) on the final assessment day (n=35). CLDA estimated the effect of time as a post-treatment BDI-II score mean of M=13.39, SE=2.74, df=75, t=4.89, p<.001, 95% CI [7.93,18.85], with a strong effect size of Cohen’s d=1.13. Contrary to the hypothesis, however, the analysis demonstrated no effect of intervention group on the primary outcome measure. The cLDA model revealed a difference in mean of diff=.07 points on the BDI-II (SE=3.17, df=75, t=.02, p=.984, 95% CI [-6.26,6.3], Cohen’s d=.00) in the psychological intervention group (n=16; M=15.75, SD=9.75) compared with the rtfMRI neurofeedback group (n=19; M=15.68, SD=10.02). Thus, the psychological intervention was shown to be of equal effect in reducing depressive symptoms compared with the rtfMRI neurofeedback training as assessed with the BDI-II (Figure 2).

**Figure 1.**
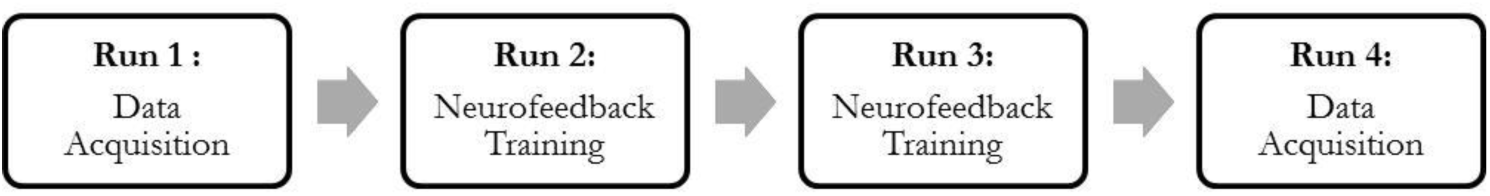
fMRI session design.

**Figure 2.**
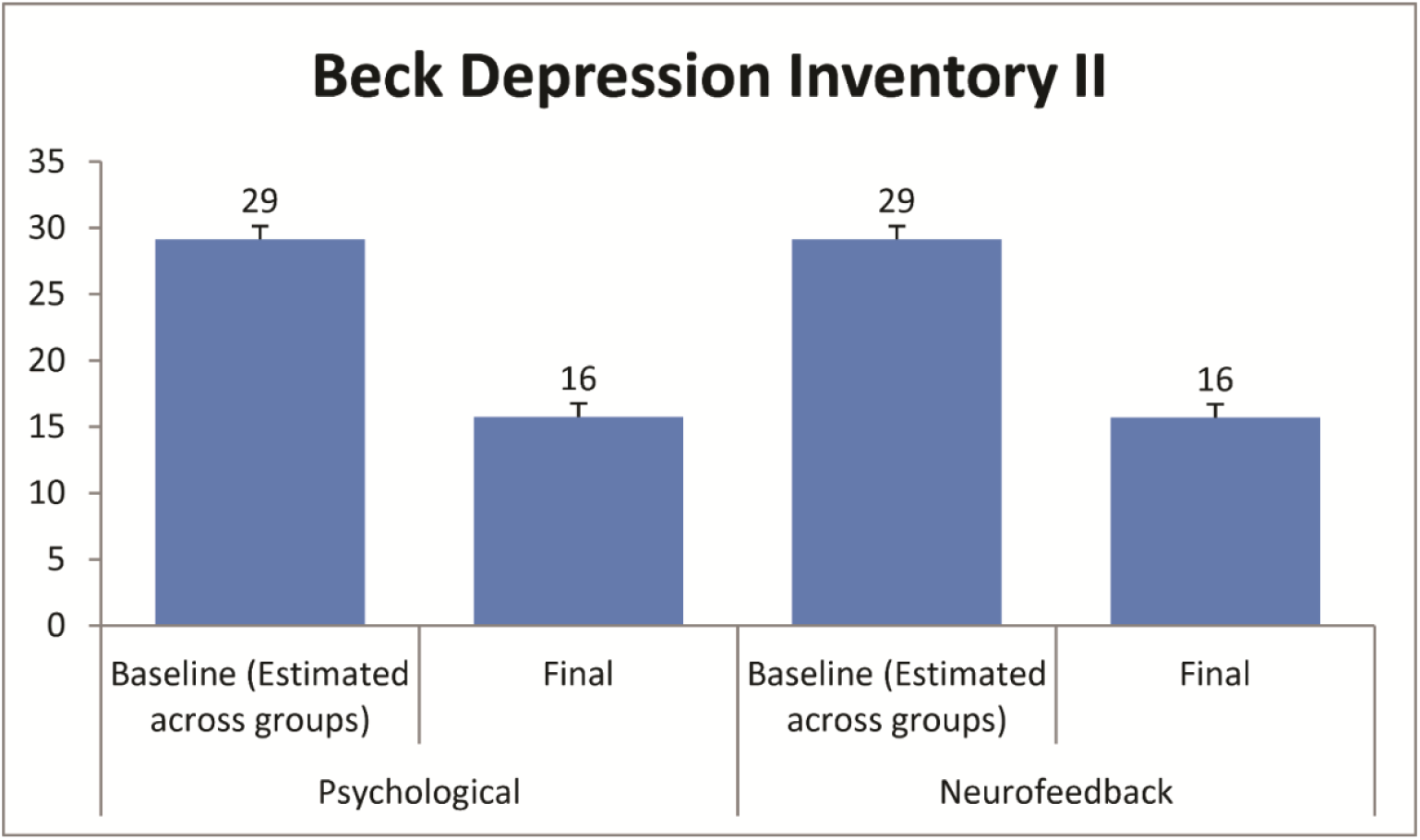
Pre- vs post-treatment comparison in score means on the BDI-II for the psychological intervention (n=16) and rtfMRI neurofeedback training group (n=19). The cLDA model estimates a common baseline BDI-II score across treatment groups (N=43). Both interventions were shown to be of equal effect with 56.25% treatment responders in the psychological intervention group vs 57.89% treatment responders in the rtfMRI neurofeedback group. Treatment response was defined as an improvement of ≥50% on the defined outcome measure.

### Pre-registered secondary outcome measures

#### Measures of depression, self-esteem and self-blame

Intervention group comparisons on pre-registered secondary outcome measures are presented in Tables 3, 4 and 5. In both intervention groups, there were significant improvements on measures of depressive symptoms, including the QUIDS-SR16 and the depression-dejection subscale of the POMS. Similarly, participants of both intervention groups showed a substantial reduction in symptoms on the MADRS on trial completion compared with baseline. Clinical global impression scales indicated a median of 2 in both groups when judged by the blinded observer (i.e. much improved) and 2.5 when self-rated in the Psychological Intervention Group and 2 in the Neurofeedback group. Moreover, MDD patients’ self-esteem increased significantly post- vs pre-treatment, regardless of the intervention group they had been allocated to. There was a clear reduction in self-blaming emotions in both groups based on the autobiographical memory ratings and when assessed by the blinded rater using a semi-structured interview designed to assess these emotions (Table 5; Zahn et al., 2015b). Nevertheless, and inconsistent with the a priori hypothesis, the rtfMRI neurofeedback intervention was not found to be superior over the psychological intervention on any of the pre-registered secondary outcome measures. Interestingly, there was also a change in implicit biases for contempt relative to anger, but none for contempt relative to anxiety, such that after the intervention, particularly in the solely psychological group, there was a reduced self-contempt bias relative to self-anger. This is in keeping with research suggesting that anger can be more adaptive as it is associated with the attribution of being able to change something and tackle it than contempt which has been associated with stable attributions of not being able to change someone’s character (Fischer & Roseman, 2007).

**Table 1.** Clinical characteristics of intervention groups

|  | PSYCHOLOGICAL<br>(n=16) | NEUROFEEDBACK<br>(n=19) |
| --- | --- | --- |
| Number of previous MDEs (percentiles) | 25<sup>th</sup>=2 50<sup>th</sup>=4.5,<br>75<sup>th</sup>=13.75<br>range: 2-66 | 25<sup>th</sup>=3 50<sup>th</sup>=4,<br>75<sup>th</sup>=8<br>range: 1-110 |
| Current MDE | 9 | 10 |
| Partially remitted | 7 | 9 |
| **MDD DSM-5 subtype** |  |  |
| Anxious Distress | 4 | 11 |
| Melancholic Features | 1 | 2 |
| Melancholic Features + Anxious Distress | 3 | 2 |
| Atypical Features | 0 | 1 |
| Atypical Features + Anxious Distress | 1 | 0 |
| None | 7 | 3 |
| **Current medication** |  |  |
| Psychotropic medication | 10 | 10 |
| Antidepressant (therapeutic dose) | 9 | 9 |
| Of which SSRI | 6 | 6 |
| **Life-time co-morbidity** |  |  |
| Current Persistent Depressive Disorder of the dysthymic subtype | 2 | 3 |
| Past PTSD with residual symptoms | 2 | 3 |
| Past PTSD fully remitted | 0 | 1 |
| Current Social Anxiety Disorder | 1 | 2 |
| Past Social Anxiety Disorder | 0 | 2 |
| Past Anorexia Nervosa | 0 | 1 |

Participants in the psychological intervention and rtfMRI neurofeedback groups did not differ on median numbers of previous episodes despite higher percentiles in the psychological intervention group ( $U=145$ , $p=.832$ , two-tailed). MDE = major depressive episode, MDD = major depressive disorder, SSRI = selective serotonin reuptake inhibitor, SNRI = serotonin-norepinephrine reuptake inhibitor, PTSD = posttraumatic stress disorder; M = mean, SD = standard deviation, [M-M] = Minimum-Maximum. In the rtfMRI neurofeedback group, one participant was suspected of displaying symptoms of autism spectrum disorder; one participant showed symptoms of attention deficit hyperactivity disorder during childhood, one participant reported past heavy alcohol and substance use.

**Table 2.** Demographic characteristics of intervention groups

|  | **PSYCHOLOGICAL** | **NEUROFEEDBACK** |
| --- | --- | --- |
|  | **M;SD;[Min-Max]** | **M;SD;[Min-Max]** |
| Age in years | 37.63;9.74;[22-55] | 36.74;11.04;[20-59] |
| Years of education | 18.06;2.52;[13-22] | 16.95;3.15;[11-23] |

Participants of both treatment groups did not differ in age ( $t=-2.50$ , $df=33$ , $p=.804$ ) or years of education ( $t=-1.14$ , $df=33$ , $p=.262$ ). In the psychological intervention group, 17/22 were female (77%) and 17/21 in the neurofeedback group (81%, Contingency Coefficient=.05, $p=.77$ , $n=43$ ).

**Table 3.**
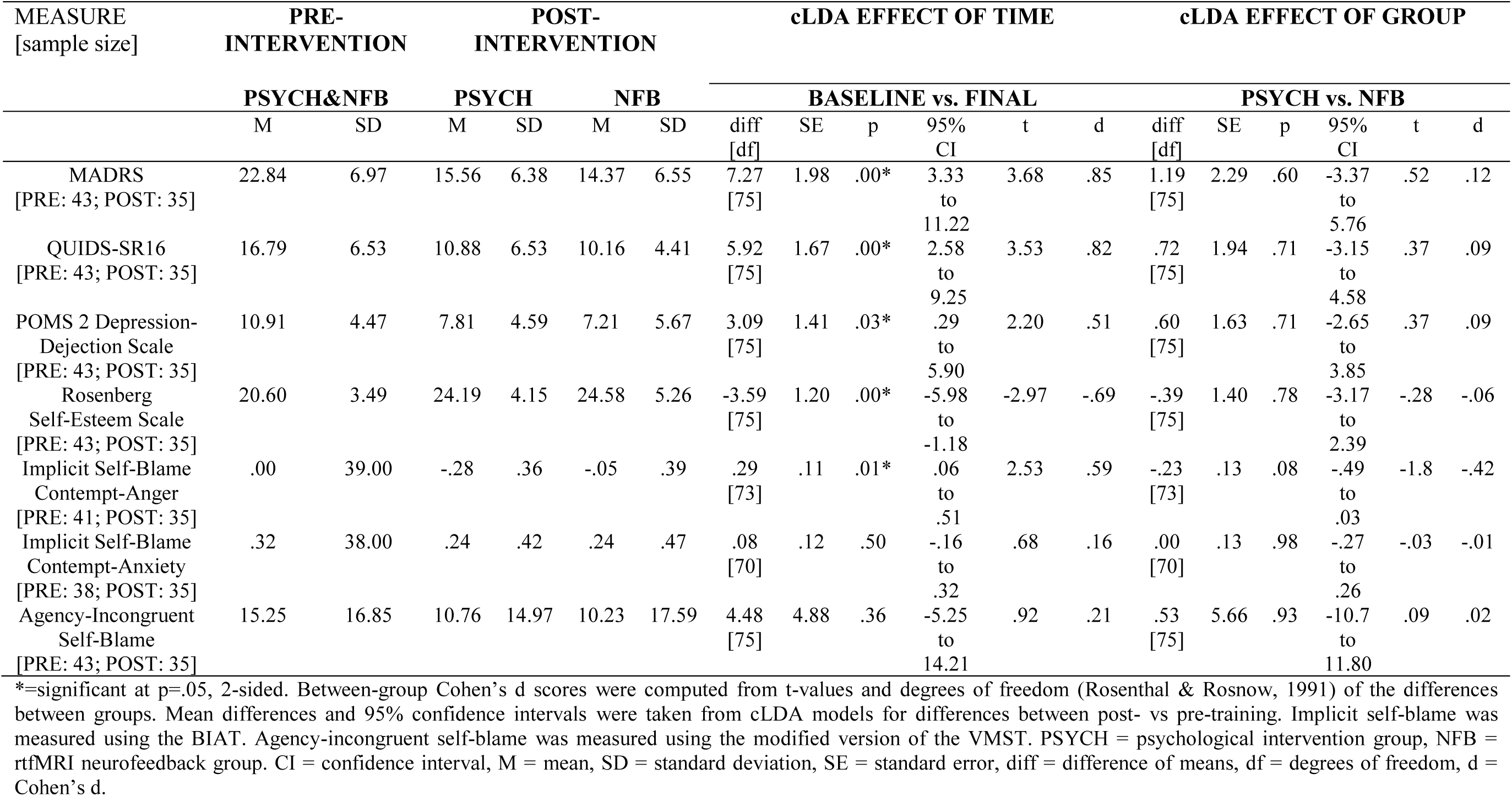
Intervention group comparisons on pre-registered secondary outcome measures

**Table 4.** Intervention group comparisons on pre-registered secondary outcome measures (cont.)

| MEASURE [sample<br>size] | MANN-<br>WHITNEY<br>U | ASYMP.<br>SIG. | EXACT<br>SIG. | MEDIAN |  | RANGE |  | MINIMUM |  | MAXIMUM |  |
| --- | --- | --- | --- | --- | --- | --- | --- | --- | --- | --- | --- |
|  |  |  |  | PSYCH | NFB | PSYCH | NFB | PSYCH | NFB | PSYCH | NFB |
| Observer-Rated CGI<br>Post-Intervention<br>[35; NFB: 19; PSYCH: 16] | 129.00 | .376 | .461 | 2.00 | 2.00 | 2.00 | 3.00 | 2.00 | 2.00 | 4.00 | 5.00 |
| Participant-Rated CGI<br>Post-Intervention<br>[35; NFB: 19; PSYCH: 16] | 139.50 | .658 | .683 | 2.50 | 2.00 | 4.00 | 4.00 | 1.00 | 1.00 | 5.00 | 5.00 |
| Withdrawal Rates<br>throughout trial<br>[43; NFB: 22; PSYCH: 21] | 218.50 | .635 |  | 0.00 | 0.00 | 1.00 | 1.00 | 0.00 | 0.00 | 1.00 | 1.00 |
| Withdrawal Rates after<br>first treatment session<br>[43; NFB: 22; PSYCH: 21] | 221.00 | .582 |  | 0.00 | 0.00 | 1.00 | 1.00 | 0.00 | 0.00 | 1.00 | 1.00 |
| Adverse Events<br>throughout trial<br>[43; NFB: 22; PSYCH: 21] | 208.00 | .352 |  | 0.00 | 0.00 | 1.00 | 1.00 | 0.00 | 0.00 | 1.00 | 1.00 |
| Adverse Events after first<br>treatment session<br>[43; NFB: 22; PSYCH: 21] | 219.50 | .527 |  | 0.00 | 0.00 | 1.00 | 1.00 | 0.00 | 0.00 | 1.00 | 1.00 |

Asymp. Sig.= 2-tailed; Exact Sig.= 2\*(1-tailed Sig.). PSYCH = psychological intervention group, NFB = rtfMRI neurofeedback training group.

**Table 5.**
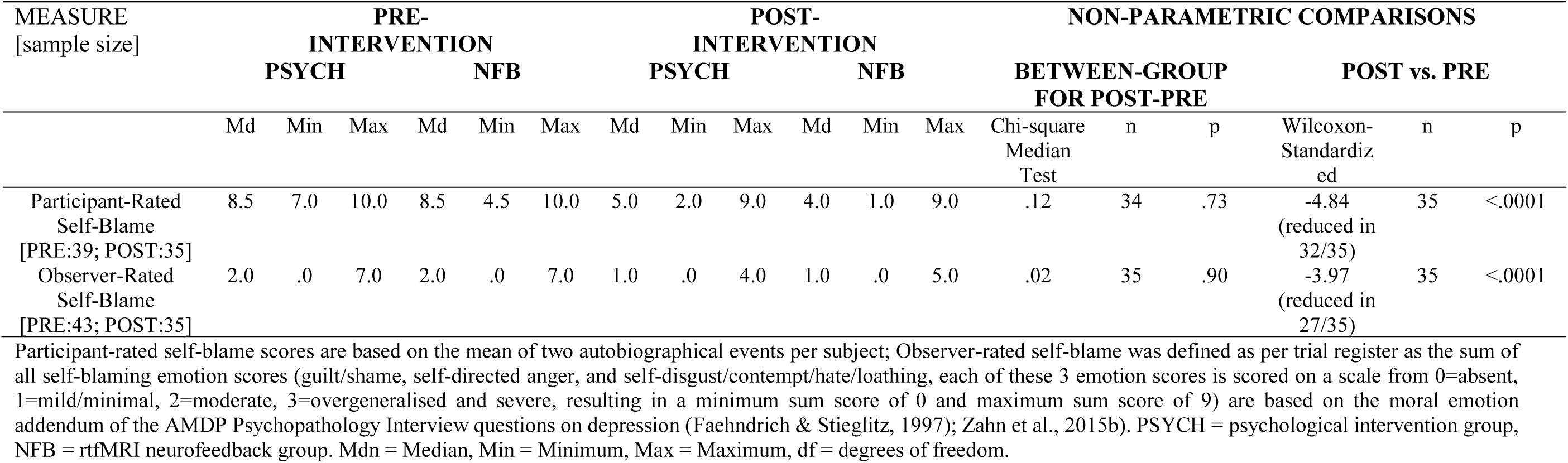
Intervention group comparisons on pre-registered secondary outcome measures (cont.)

### Adverse events and withdrawal rates

Adverse event and withdrawal rates were pre-registered as additional secondary outcome measures. Notably, rtfMRI neurofeedback training as well as the solely psychological treatment were found to be safe and feasible forms of intervention. No significant differences emerged regarding adverse events or withdrawal rates throughout the trial following randomisation or specifically, between the first treatment session and trial completion (Table 4). Overall, six adverse events were reported following randomisation, three occurring at a time point subsequent to the first treatment session. A possible relationship with the study has been suspected in four of the overall six adverse events throughout the trial, while no relationship was observed in one case and a probable relationship was assumed in one other instance. In the latter case, the participant reported transient insomnia lasting one night after his first rtfMRI neurofeedback session.

All adverse events were mild and constituted of two withdrawals and one incident of exclusion from the study. The participant was excluded on the day of the first treatment session, after having been randomised to the psychological intervention group, as the patient presented with symptoms of depression that had worsened by 10 points on the BDI-II between baseline assessment and first intervention day. In addition, one adverse event occurred prior to randomisation and had no relation with the study. The participant’s result on the ACE-III was suggestive of a neurological condition, and further referral to a cognitive assessment service was recommended.

Throughout the trial, seven participants withdrew their consent and ended their participation in the study, four prior to the first day of intervention and three at different time points following their first treatment session. In the former instance, participants reported to not feel well enough to participate or to experience time-related challenges that would make it impossible to attend the five scheduled study appointments. In the situation of participants withdrawing after their first treatment session, they described family or financial reasons for their decision. As aforementioned, transient insomnia in the night following the first rtfMRI neurofeedback session was cause for one MDD patient to discontinue trial participation.

### Real-time fMRI neurofeedback group: rSATL–posterior SC connectivity post- vs pre-intervention

As a further secondary outcome measure, specific to the rtfMRI neurofeedback intervention group, functional connectivity between the rSATL and the posterior SC was measured for self-blame relative to blaming others, post- vs pre-intervention. Change in functional connectivity was assessed by calculating Cohen’s d effect sizes for regression coefficient means for time series pre- and post-rtfMRI neurofeedback training. As predicted, a significant training-induced reduction in connectivity between the rSATL and posterior SC was detected in the guilt condition relative to indignation, as reflected in a significant time x condition interaction in a repeated measures ANOVA (Table 6). Inconsistent with the prediction, this decrease was not found to be significant for the guilt condition itself (t=-.89, df=17, p=.387; n=18), the mean difference between conditions was - 1.27 with a 95% confidence interval between -.43 and .18. Interestingly, as guilt-specific connectivity successfully reduced relative to indignation post-treatment, indignation-related connectivity between the rSATL and posterior SC was observed to increase with a mean difference of .09 post- vs pre-rtfMRI neurofeedback training (Figure 3; Table 6). This finding, however, was not significant itself (t=.68, df=17, p=.504, 95% CI [-1.76,3.43].

**Table 6.** Post-training vs pre-training comparison of pre-registered secondary outcome measure rSATL – posterior SC connectivity on fMRI

| MEASURE [sample size] | PRE-TRAINING |  | POST-TRAINING |  | REPEATED MEASURES ANOVA |  |  |  |  |  |
| --- | --- | --- | --- | --- | --- | --- | --- | --- | --- | --- |
|  | NFB |  | NFB |  | TIME |  | CONDITION |  | TIMExCONDITION |  |
|  | M | SD | M | SD | F | p | F | P | F | p |
| Guilt: rSATL-SC regression<br>coefficient: Cohen's d | .29 | .54 | .16 | .37 | .29 | .87 | .43 | .52 | 6.40 | .02* |
| Indignation: rSATL-SC<br>regression coefficient: Cohen's d | .15 | .46 | .24 | .25 |  |  |  |  |  |  |

\*=significant at p=.05, 2-sided. NFB = rtfMRI neurofeedback group, rSATL-SC = right superior anterior temporal lobe - posterior subgenual cortex. n=18 with complete data.

**Figure 3.**
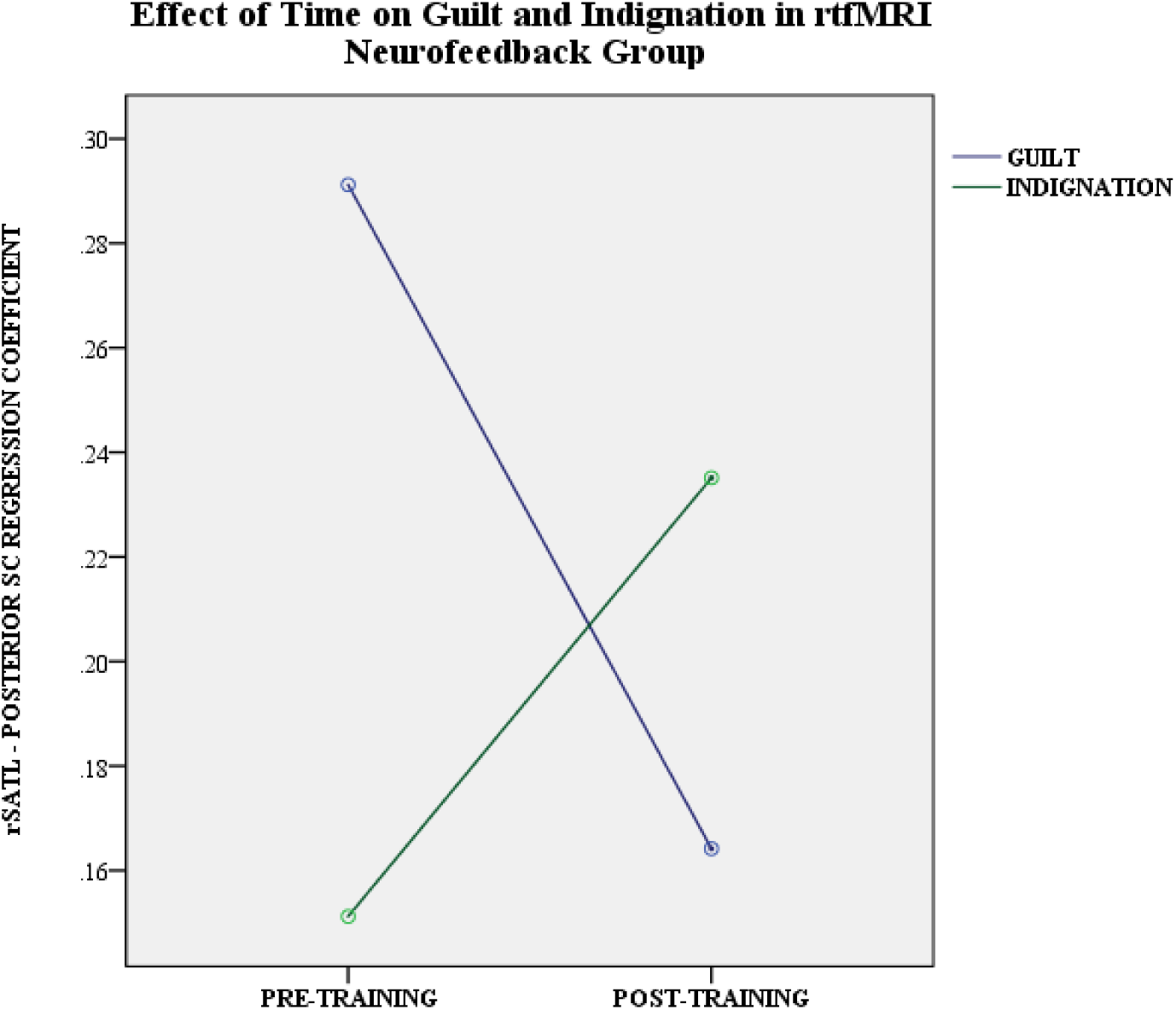
Relative change in functional connectivity between rSATL and posterior SC in the guilt and indignation condition, measured as Cohen’s D for regression coefficient means for time series pre- and post-rtfMRI neurofeedback training, comparing the first and final treatment session.

In addition to applying the CLDA model, the intention-to-treat (ITT) approach was chosen and compared with the per-protocol analyses, using the Pearson Chi-Square test to analyse the association between intervention group and treatment response on the primary outcome measure (BDI-II). The ITT analysis includes data of all randomised participants regardless of their adherence or withdrawal subsequent to randomisation (Fisher et al., 1990). Here, participants who withdrew from the study or did not complete the trial were treated as non-responders. No relationship was found between intervention group and treatment response χ2(1, N=43)=.029, p=.864. The estimated treatment effect is considered to be conservative in ITT analysis and caution is raised in terms of ITT’s susceptibility to type II errors (for a review see Gupta, 2011). Contrary to the ITT analysis, the per-protocol analysis risks to falsely present a treatment effect (type I error). It excludes participants who withdraw after randomisation and disregards data of those who do not complete the study. The per-protocol analysis may lead to significant reductions in statistical power by affecting the overall sample size (Gupta, 2011). Using this analysis, no association between intervention group and treatment response was found χ2(1, N=35)=.046, p=.830. The results of these additional analyses confirm findings based on the cLDA model and were contrary to our predictions.

Throughout all treatment sessions and active neurofeedback runs, participants were able to successfully bring down the level of guilt-associated correlations reflected in an average neurofeedback thermometer level around ∼50% (Supplementary Figure 4). This is remarkable considering that FRIEND implements a moving target algorithm potentially making it more difficult to control the neurofeedback thermometer as connectivity between the rSATL-posterior SC successively reduces with training.

A repeated measures ANOVA was conducted to investigate differences in rSATL-posterior SC functional connectivity pre- vs post-intervention in the guilt vs indignation condition over the course of all three treatment sessions. This analysis approach was chosen to contrast the two psychological conditions and thereby control for non-specific correlations, which make up a large fraction of the signal when considering each condition in isolation. While a significant main effect was found for pre- vs post-intervention (F(1,17)=4.5, p=.049, Wilks’ Lambda=.79, ηp^2^=.21), there was only a trendwise interaction between session and pre- vs post-interventional rSATL-posterior SC connectivity (F(2,16)=2.79, p=.091, Wilks’ Lamda=.74, ηp^2^=.26). Pre- and post-interventional connectivity measures and their change over the course of all three neurofeedback training sessions are displayed in Supplementary Figure 5. This shows that guilt connectivity was indeed reduced after the neurofeedback training relative to indignation in concordance with our main analysis. It appears that most of this training effect occurred already after the first session (Session A), but this observation was only supported by a trendwise interaction between session and intervention effect.

### Exploratory secondary data analysis

#### Major depressive disorder with and without anxious distress

Based on the primary finding of equally strong treatment responses in both intervention groups, subsequent exploratory data analyses of clinical subtypes of MDD investigated differences in the response of MDD patients with and without anxious distress. A univariate GLM showed a significant interaction between treatment group and anxious distress features (F(1,30)=4.98, p=.033, ηp^2^=.14). This interaction was due to a better response to the neurofeedback-enhanced intervention in non-anxious (83% of patients halving their BDI-II scores) vs. anxious patients (46%) and a better response to the solely psychological intervention in anxious (75%) vs. non-anxious (38%) patients. There was no significant main effect of the MDD anxious/non-anxious subtype (F(1,30)=.78, p=.782, ηp^2^ =.003), nor a main effect of treatment group (F(1,30)=0, p=.989, ηp^2^ =0; Figure 4).

**Figure 4.**
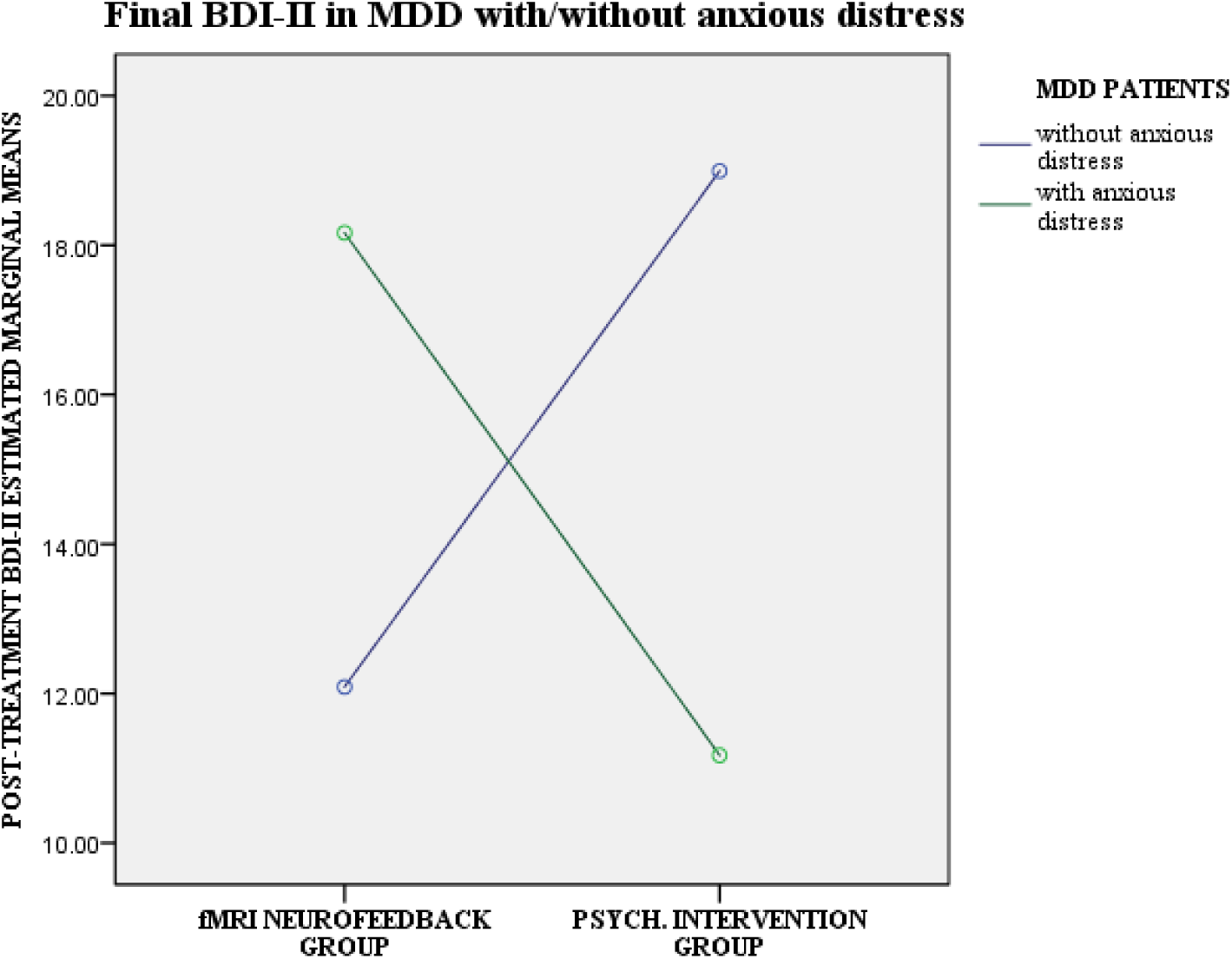
The results of a secondary analysis are displayed which stratified our primary outcome by anxious distress features, the most frequent major depressive disorder (MDD) subtype in our trial (n=21 out of n=35, Structured Clinical Interview for DSM5). Plotted are post-treatment BDI-II estimated marginal means of MDD patients with and without anxious distress in both treatment groups (fMRI neurofeedback group: n=19; psychological intervention group: n=16). Covariates appearing in the model are evaluated at the estimated baseline BDI-II value of 28.6 points.

In Supplementary Table 2, we also explored baseline differences between randomised anxious and non-anxious distress subtype MDD patients on clinical and demographic measures to determine whether our stratified analysis could have been confounded by other clinical differences. There was no difference on demographic variables, number of previous episodes, or baseline BDI-II scores, or current effective antidepressant medication. Between completers, there was also no difference in engagement with the therapeutic strategies. As one would predict from the association of anxiety and irritability/anger in DSM-5 diagnostic criteria for anxiety disorders such as generalised anxiety disorder and posttraumatic stress disorder, we found higher levels of current anger towards others in the anxious compared with the non-anxious MDD group as measured on our psychopathological interview. Interestingly, there was no subtype difference in the sum of self-blaming emotions score computed as a secondary outcome measure using this interview. There were only 8 of 43 patients (19%) who did not experience at least one self-blaming emotion (self-directed anger, self-contempt/disgust, shame or guilt, or a combination of shame and guilt) to a bothering degree, but this absence of self-blaming emotions was not associated with anxious distress, on the contrary, there was a trend towards anxious distress patients showing more consistent self-blaming emotions on our interview. This indicates that both self- and other-blaming emotions were prominent in the anxious distress subtype.

#### Change in connectivity on fMRI, self-esteem and engagement in treatment

Based on previous findings, where measures of self-esteem correlated with changes in functional connectivity between the rSATL and the anterior subgenual cingulate after rtfMRI neurofeedback training (Zahn et al., 2018), non-parametric correlation analyses were conducted to explore this pattern in the rtfMRI neurofeedback group (Table 7). Notably, no such correlation was found, a change in connectivity between the rSATL and the posterior SC in guilt relative to indignation was not associated with an increase in self-esteem in the patient group. Interestingly, however, improvement in depression scores correlated with an increase in self-esteem. Similarly, a positive correlation was found between increased self-esteem and engagement in treatment as assessed by the summed frequency of use of treatment-specific psychological strategies throughout the study in both intervention groups.

**Table 7.**
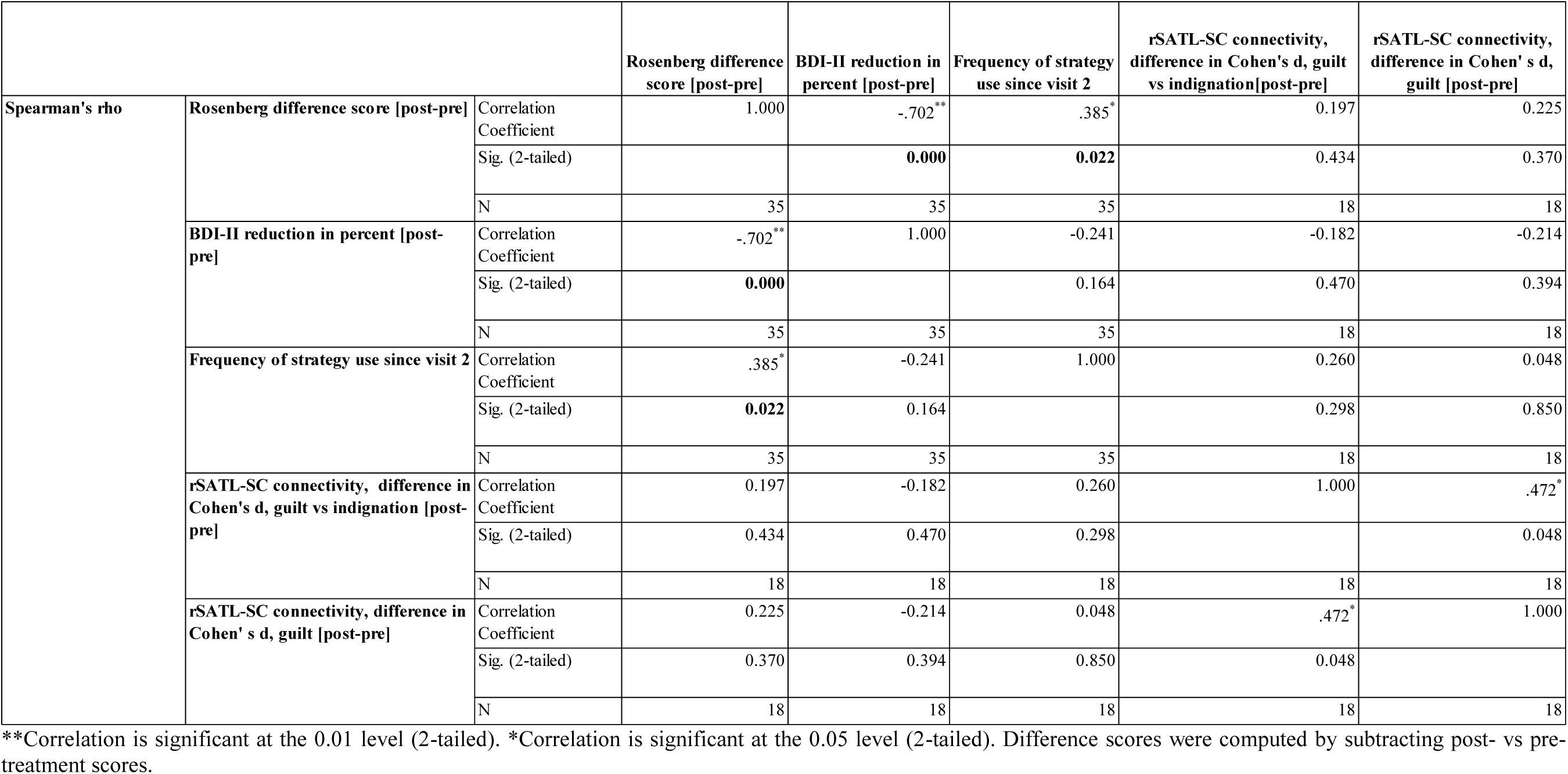
Secondary correlation analyses

## Discussion

### Discussion of main findings

This single-blind, randomised, controlled pilot trial investigated the clinical potential of a novel rtfMRI neurofeedback protocol compared to the therapeutic effects of a newly designed self-guided psychological intervention in early treatment-resistant MDD. It was hypothesised that patients randomised to the rtfMRI neurofeedback group show a reduction in depressive symptoms and self-blame while exhibiting an increase in self-worth compared to the psychological intervention group. Furthermore, it was proposed that patients undergoing rtfMRI neurofeedback training show a decreased functional connectivity between the rSATL and the posterior SC post-treatment compared to pre-treatment. Decreased functional connectivity between the rSATL and posterior SC region was predicted to be associated with a reduction in depressive symptoms in the rtfMRI neurofeedback group.

The results demonstrated that both interventions were safe, with no relevant adverse events occurring in either group. There was a strong effect size for patients’ improvement on self-rated and observer-rated depression measures with response rates above 55% in both intervention groups. Thus, the safety and overall clinical benefits of the rtfMRI neurofeedback intervention in MDD is in keeping with previous studies (Linden et al., 2012, Mehler et al., 2018, Young et al., 2014, Young et al., 2017a). This is particularly remarkable as the NeuroMooD protocol asked participants to engage with negative rather than positive emotions, opposed to previous studies (Linden et al., 2012, Mehler et al., 2018, Young et al., 2014, Young et al., 2017a). Contrary to the first hypothesis, no difference was found between the rtfMRI neurofeedback and the psychological intervention group on the primary outcome measure (BDI-II). The second prediction was confirmed as the rtfMRI neurofeedback training resulted in a decrease in functional connectivity between the rSATL and the posterior SC for guilt relative to indignation. Contrary to the third hypothesis, no relationship was found between connectivity changes and the changes in depressive symptoms after three sessions of rtfMRI neurofeedback training.

Various considerations need to be taken into account as to why no group differences were found on primary and secondary outcome measures. One possibility might be that the improvements observed in both intervention groups were due to spontaneous remission or placebo-like effects instead of being the result of the experimental treatment. This is possible, yet unlikely to be the only explanatory factor for this finding as the placebo response rate in MDD is generally found to be lower, usually around 30% (Walsh, Seidman, Sysko &Gould, 2002), well below the >55% response rate demonstrated in both treatment groups in the NeuroMooD trial. Furthermore, the NeuroMooD protocol aimed at minimising the risk of including spontaneously remitting patients by including MDD patients only if they were stable, i.e. with no improvement, in symptoms for at least six weeks before randomisation into the study, and by restricting inclusion to patients with early treatment-resistance and recurrent MDD. Lastly, the frequency of how often participants used the psychological strategies between treatment visits was found to positively correlate with an increase in self-esteem in both intervention groups, which further argues against spontaneous remission as the sole explanation for the observed findings. Another possible explanation why rtfMRI neurofeedback did not show superiority over the psychological intervention might be because the rtfMRI neurofeedback intervention provided no added value in reducing symptoms of depression in early treatment-resistant MDD compared with the strong effects of the self-guided psychological intervention. While this explanation cannot be ruled out, the secondary data analysis suggests that rtfMRI neurofeedback training is superior to the psychological intervention in the non-anxious distress subtype of MDD. This finding will, however, require further confirmation in future studies.

The observed rtfMRI neurofeedback training-induced reduction in functional connectivity between the rSATL and the posterior SC for guilt relative to indignation demonstrates that MDD patients were able to successfully modulate their brain connectivity as guided by the rtfMRI neurofeedback signal. The lack of association between functional connectivity changes and improvement in the severity of depressive symptoms is in keeping with the limited clinical benefit in the rtfMRI neurofeedback group overall. Considering that the majority of MDD patients in the rtfMRI neurofeedback group were of the anxious distress subtype, the neural fMRI target may be irrelevant for the anxious distress subtype of MDD; a hypothesis that needs to be examined in future larger studies.

## Limitations

The following potential limitations of the NeuroMooD trial need to be considered: firstly, the study might have been underpowered and, therefore, unable to detect a clinically meaningful difference between the two intervention groups. Nevertheless, the effect sizes for non-superiority of the rtfMRI neurofeedback group were so small, that even a large sample would have been unable to find differences between groups. Furthermore, the trial’s sample size was comparable to other randomised clinical trials investigating rtfMRI in MDD (Mehler et al., 2018; Young et al., 2017a). Our secondary data analyses stratifying for subtype were limited by not being pre-registered and by the relative scarcity of non-anxious MDD patients and thus need reproducing in a larger sample.

Contrary to previous rtfMRI neurofeedback studies in MDD (Young et al., 2014, Young et al., 2017a) that included medication-free patients, around half of MDD patients in each group participating in the NeuroMooD trial were taking antidepressant medication. Despite the possibility that antidepressant medication may have negatively impacted on the participants’ performance during the rtfMRI neurofeedback training, Linden et al.’s, (2012) pioneering rtfMRI neurofeedback study targeted a sample on stable antidepressant medication comparable to NeuroMooD and demonstrated the superiority of the rtfMRI neurofeedback relative to the control condition. Finally, and opposite to Young et al. (2017a), the NeuroMooD study was limited by lacking a rtfMRI neurofeedback control arm, which was deliberate in order to probe its clinical usefulness against a cheaper intervention. Had we found rtfMRI superiority over the control intervention, this would have led to some difficulties in ruling out non-specific placebo-like effects of neurofeedback. The fact that there was no superiority of rtfMRI overall in our study, suggests that being in a scanner environment itself did not have strong Placebo-like effects. Furthermore, control neurofeedback interventions are difficult to design and interpret. Young et al. (2017a) used the left intraparietal sulcus signal in the control neurofeedback condition which is not relevant for recalling positive emotions, the task given to participants. This mismatch between neurofeedback signal and psychological instructions could have contributed to the inferiority of the control intervention.

## Conclusion

Both NeuroMooD interventions were demonstrated to be safe and resulted in a reduction in symptom severity of 46% and a treatment response of more than 55% in the study sample of current and insufficiently remitted, early treatment-resistant MDD. Although a contribution of placebo-like and non-specific effects cannot be ruled out, it is likely that the self-guided psychological intervention has had additional specific therapeutic effects. Our secondary data analysis suggests that self-blame-selective rtfMRI neurofeedback training is of superior benefit in non-anxious MDD patients compared with the solely psychological intervention, which needs further confirmation in a larger sample.

## Data Availability

The datasets generated during and/or analysed during the current study are/will be available upon request from Dr Roland Zahn. No personal data will be shared and the data is fully anonymised. The data management plan stipulates access:
5.1. Suitability for sharing
The anonymised data are suitable for sharing with collaborators.
5.2. Discovery by potential users of the research data
Potential users are pointed to the data through publications and the study website and can apply for data sharing via a collaboration. They will be able to get access to the data directly through data repositories such as the one recently launched at KCL with searchable metadata (www.kcl.ac.uk/library/researchsupport/research-data-management/ DepositPublishPromote/Deposit-your-data-with-Kings.aspx) after an embargo period of 5 years.
5.3. Governance of access
Primary decisions will be made by the PI, and subsequently reviewed by an independent advisor.
5.4. The study team’s exclusive use of the data
There will be an embargo of 5 years after study completion during which only the original research team and new researchers in the PI’s lab or invited collaborators will have access.
5.5. Restrictions or delays to sharing, with planned actions to limit such restrictions
See above
5.6. Regulation of responsibilities of users
People outside the research team who want to collaborate and use data will have to agree to a data sharing agreement in line with MRC principles.

## Acknowledgements

We are very grateful to all participants of this study and their time and effort. We are also grateful to our funders: the NARSAD independent investigator award to RZ awarded by the Brain & Behavior Research Foundation, the IoPPN at King’s College London as well as the NIHR Biomedical Research Centre for Mental Health at the South London and Maudsley NHS Foundation Trust. The views expressed are those of the author(s) and not necessarily those of the NHS, the NIHR or the Department of Health and Social Care. We are grateful to Dr Karen Lythe, Professor Galen Bodenhausen, and Professor Nicolas Rüsch for their important contribution to the design of the BIAT task.

## Supplementary Materials

### Supplementary Methods

#### Brief Implicit Association Test Design and Analysis

The self-contempt Brief Implicit Association Test (BIAT), an experimental measure developed by R.Z. and Dr Lythe in collaboration with Prof Rüsch and Prof Bodenhausen, was employed as an indirect measure of self-contempt biases, evaluating the association of contempt or disgust with oneself relative to others. Further, we included an established BIAT (Sriram & Greenwald, 2009) used to assess implicit self-esteem. The rationale for the development of this computerised task was based on the endeavour to measure self-contempt biases without the participants’ awareness of what the task captures. This strategy is meant to prevent distortions in the participants’ response. The task design is based a similar test, the Implicit Association Test (IAT; Greenwald, McGhee & Schwartz, 1998), which has been validated to measure implicit self-esteem (Greenwald & Farnham, 2000).

The BIAT uses complementary pairs of categories and attributes which the participant needs to classify. The speed in which participants respond is reflective of the strength of the association within the two pairs of categories that are paired on the same response key (Greenwald & Farnham, 2000; Greenwald, McGhee, & Schwartz, 1998). Specifically, participants are instructed to decide with a response if two sets of items match an associated category-attribute pair and are asked to give a different response should the item pairs not match (Greenwald & Farnham, 2000). Participants respond as quickly as possible by pressing left and right keys on a computer keyboard (Greenwald & Farnham, 2000). As aforementioned, the automatic association between a category (e.g. self) and an attribute (e.g. contemptuous) is assessed by calculating differences in speed between two conditions. In condition 1, words indicative of ’self’, i.e. the participant, and words relating to contempt share the same response key (i.e. require the pressing of the right-side key). In condition 2, words that address the ’self’ (i.e. the participant) and words that relate to anxiety share the same response key (here, the left key needs to be pressed).

The BIAT consists of two experimental tasks that assess implicit self-contempt, whereby the categories consist of self-agency, other-agency, contempt and a non-focal category which is either anxiety or anger. In the implicit self-esteem task, the categories ’self’ and ’other’ are attributed to ’good’ (positive valence) and ’bad’ (negative valence). Greenwald et al. (2002) emphasise that this task design, due to its use of complementary pairs of concepts and attributes, is limited to measuring the relative strength of pairs of associations rather than the absolute strength of single associations. The authors conclude, however, that the task is meaningful in practice due to the opposing, yet complementary quality of many socially meaningful categories, i.e. good and bad (Greenwald et al., 2002; Greenwald & Farnham, 2000).

BIAT data was exported from Inquisit 3 (https://www.millisecond.com/products/Inquisit3/) and analysed with SPSS 24 (https://www.ibm.com/analytics/spss-statistics-software). Scoring algorithms and analyses strategies were based on the improved scoring algorithm created by Nosek (2005, May 27). Trials with an error rate of higher 30% of 32 trials were excluded from the analyses; similarly, trials, where more than 10% of participant responses had a latency of less than 300 milliseconds, were excluded. Self-contempt bias was measured by subtracting the mean value of latency for the category ’self and contempt’ (16 trials) minus the mean value of the latency for the category ’other and contempt’ (16 trials), divided by the standard deviation of latency computed for all 32 trials. Hereby, a more positive total score is understood as being indicative of a higher degree of self-contempt.

The Contempt vs. Anxiety BIAT consisted of four categories: Self-agency, Other-agency, Contempt, [Anxiety] as non-focal category. In Block 1, participants pressed the left key for “Participant acts” OR “Contempt”. They pressed the right key for everything else. In Block 2, they had to press the left key for “Friend acts” OR “Contempt” and pressed the right key for everything else. Stimuli presented for categorisation were: “Participant acts”, “Participant does”, “Participant makes”, “Participant causes” as examples of Self-agency. The same stimuli, but with the best friend’s name were used for other-agency. “Hate”, “disgust”, “contemptuous”, “contempt” were used as examples of the Contempt category. “Anxiety”, “anxious”, “fear”, “scared” were used as examples of the Anxiety category. The Contempt vs. Anger BIAT was constructed in the same way, only replacing Anxiety with the Anger category and using “anger”, “angry”, “fury”, “furious” as examples for categorisation.

### Supplementary Results

**Supplementary Table 1.** Trial schedule chart.

|  | **Initial Patient Contact** | **Pre-Trial Assessment Visit 1** | **Treatment Session Visit 2** | **Treatment Session Visit 3** | **Treatment Session Visit 4** | **Post-Trial Assessment Visit 5** |
| --- | --- | --- | --- | --- | --- | --- |
|  | [email & phone] | [day 0] | [1-13 days after visit 1] | [7-13 days after visit 2] | [7-13 days after visit 3] | [7-13 days after visit 4] |
| **Oral informed consent** | ✓ |  |  |  |  |  |
| **Introduction to the clinical trial** | ✓ |  |  |  |  |  |
| **Assessment of eligibility** | ✓ | ✓ |  |  |  |  |
| **Written informed consent** |  | ✓ | ✓ | ✓ | ✓ | ✓ |
| **Clinical & neuro-psychological assessment** |  | ✓ |  |  |  | ✓ |
| **Psychological Intervention only** |  |  | ✓ | ✓ | ✓ |  |
| **rtfMRI neurofeedback training** |  |  | ✓ | ✓ | ✓ |  |
| **Mood assessment** |  | ✓ | ✓ | ✓ | ✓ | ✓ |
| **Risk assessment** |  | ✓ | ✓ | ✓ | ✓ | ✓ |

**Supplementary Table 2.** Comparison of anxious and non-anxious distress subgroups.

| Measure | Anxious MDD | Non-anxious<br>MDD | Statistic | Significance |
| --- | --- | --- | --- | --- |
| Years of education | 17.3±2.6 | 17.3±3.4 | t=0.02 | .98 |
| Age | 38.6±10.2 | 34.2±8.4 | t=-1.4 | .16 |
| Gender | 22/28 female<br>(79%) | 12/15 female<br>(80%) | CC=.02 | .91 |
| Number of previous MDEs<br>(median) | 3.5 | 5 | Median<br>Test<br>$\chi^2=.11$ | .74 |
| Current antidepressant<br>medication at therapeutic dose (n) | 15/28 (53.6%) | 7/15 (47.7%) | CC=.07 | .67 |
| BDI-II at Baseline | 29.9±9.6 | 27.8±6.8 | t=-.7 | .46 |
| Frequency of therapeutic<br>strategy use (m) | 9.1±2.5 | 7.6±3.1 | t=-1.4 | .17 |
| AMDP Anxiety (n) | 19/28 (67.9%) | 5/15 (33.3%) | CC=.32 | .03 |
| AMDP Anger towards others (n) | 10/28 (35.7%) | 0/15 (0.0%) | CC=.37 | .008 |
| Sum of self-blaming emotions<br>score (median) | 2.5 | 2 | Median<br>Test<br>$\chi^2=.09$ | .76 |
| Any self-blaming emotion (n) | 25/28 (89.3%) | 10/15 (66.6%) | CC=.27 | .07 |

Anxious distress features were defined on the Structured Clinical Interview for DSM5 for the current episode.
MDE = major depressive episode, MDD = major depressive disorder, BDI-II = Beck Depression Inventory-II; m = mean, ± = standard deviation, [M-M] = Minimum-Maximum. CC=Contingency Coefficient in Crosstab statistics. The frequency of therapeutic strategy use relied on completion and was collected in all n=35 completers (n=21 anxious MDD, n=14 non-anxious MDD). All other measures were collected at baseline in all randomised patients (n=43, n=28 with and n=15 without anxious distress). Baseline psychopathological measures (AMDP anxiety and anger towards others) were based on binarising scores for the past two weeks into 0=absent or mild or 1=moderate or severe as previously (Zahn et al., 2015b). The sum of self-blaming emotions score on the AMDP was based on our secondary outcome measure as described above. The any self-blaming emotion measure was based on identifying patients who had no self-blaming emotion to a moderate degree on the moral emotion addendum to the AMDP (i.e. no self-directed anger, shame, guilt, or self-disgust/contempt). Significance reflects 2-sided p-values or approximate significance for Contingency Coefficient statistics.

**Supplementary Figure 1:**
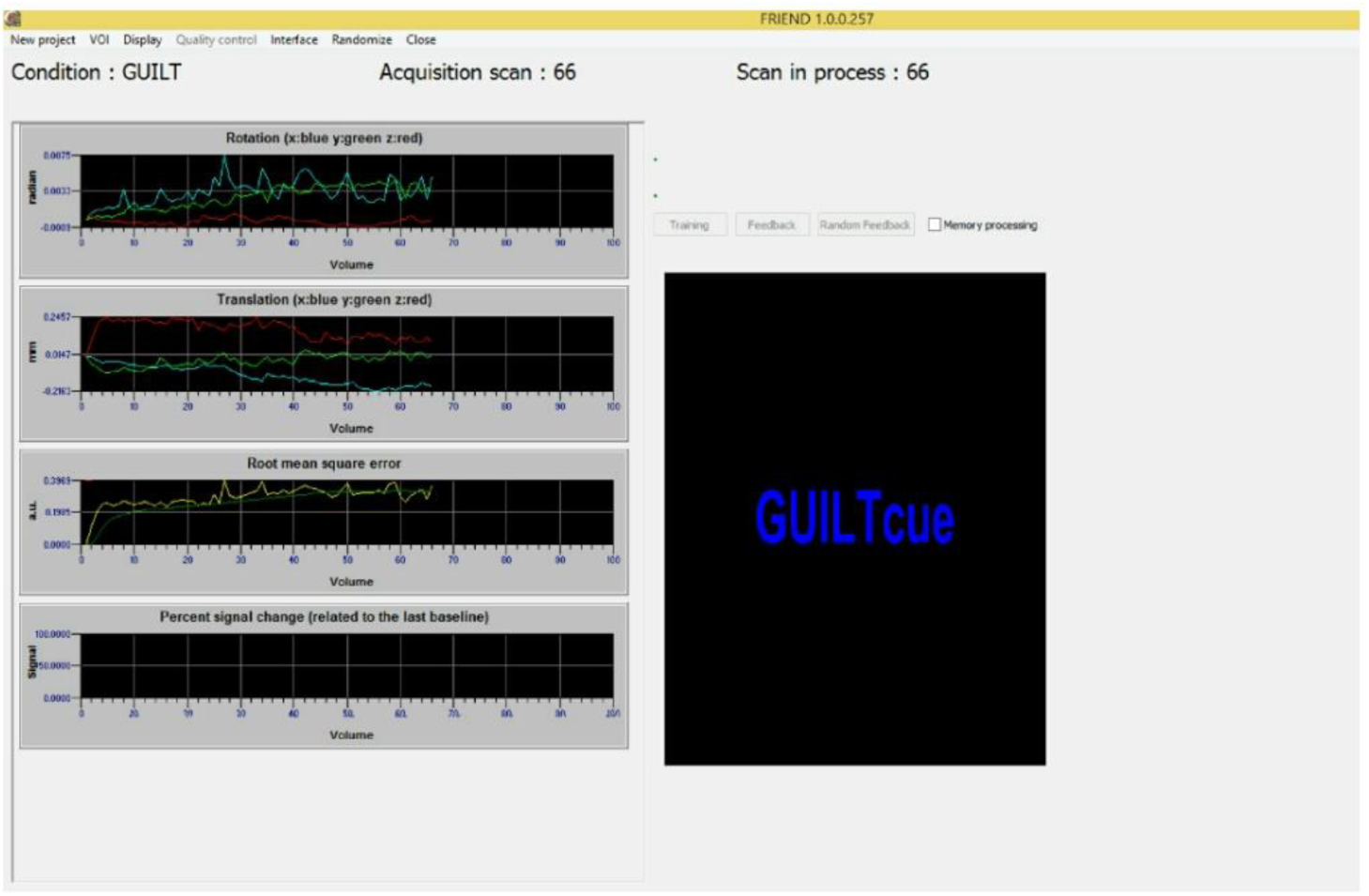
Display of the interface of the rtfMRI neurofeedback software FRIEND during a data acquisition run (Basilio et al., 2015; Sato et al., 2013). During acquisition runs (run 1 and 4), participants are presented with their guilt and indignation/anger cue words. Cue words refer to autobiographical memories of events that trigger patients to experience feelings of self-blame or indignation/anger. During these runs, participants are solely thinking about these events and are not using any psychological strategies to manage associated feelings of self-blame. In-between the emotional conditions, participants are presented with numbers which cue them to perform mental subtractions from a number displayed at the screen. The right panel displays what the participant sees in the scanner, the left panel displays motion parameters and percent signal change in the regions of interest.

**Supplementary Figure 2:**
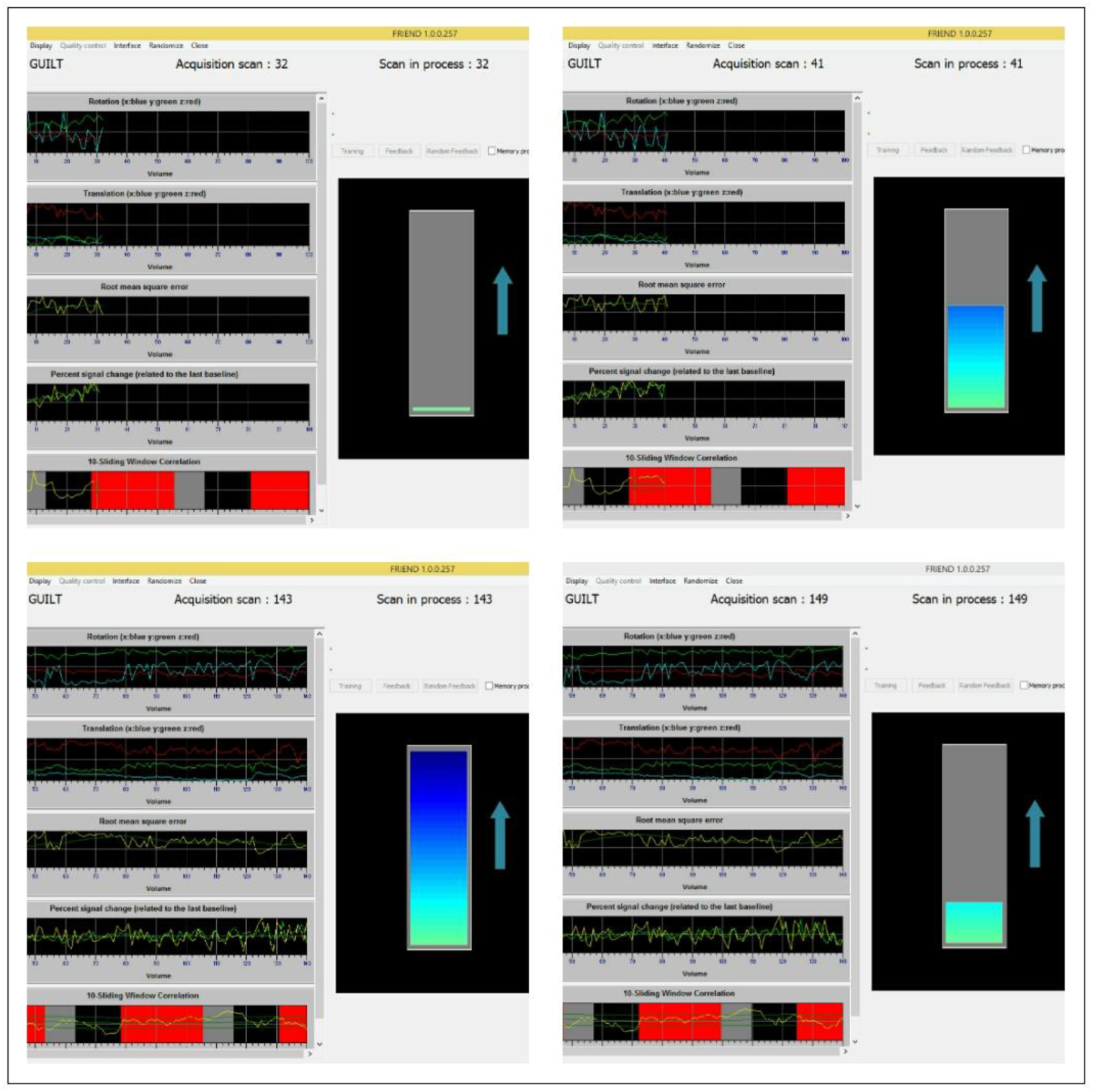
Display of the interface of the rtfMRI neurofeedback software FRIEND during a neurofeedback training run. In the scanner, participants are presented with their individualised guilt cue words or subtraction blocks. In the guilt condition, a thermometer display containing a moving colour bar appears and represents visual feedback of the patient’s functional connectivity patterns in real-time. During the rtMRI neurofeedback runs (run 2 and 3), participants are asked to bring up the level of the thermometer by using psychological strategies when thinking about autobiographical guilt-evoking events. The colour bar rises if functional (hyper-)connectivity between the rSATL and posterior SC successfully decreases.

**Supplementary Figure 3.**
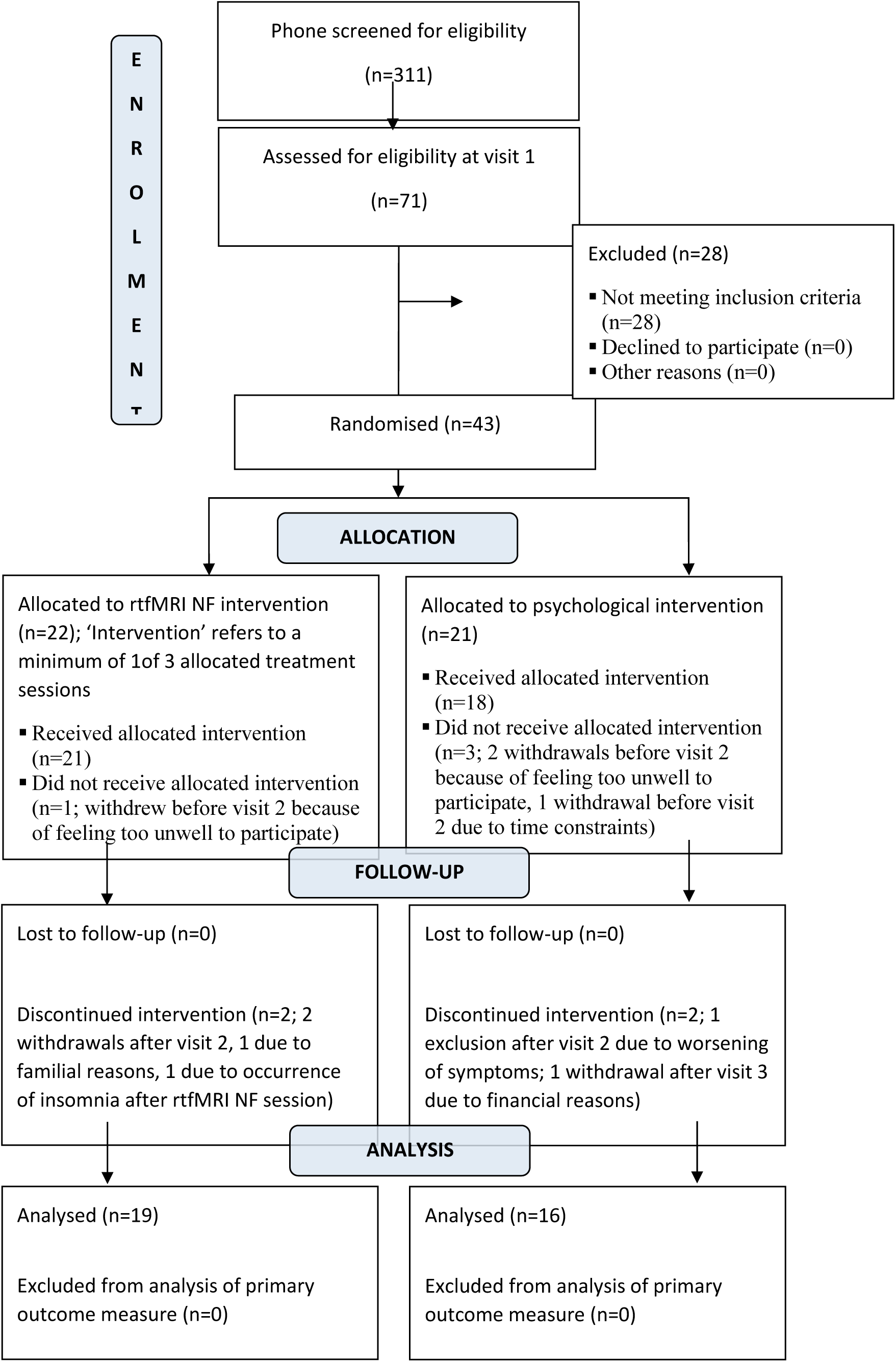
Consort Trial flow diagram.

**Supplementary Figure 4.**
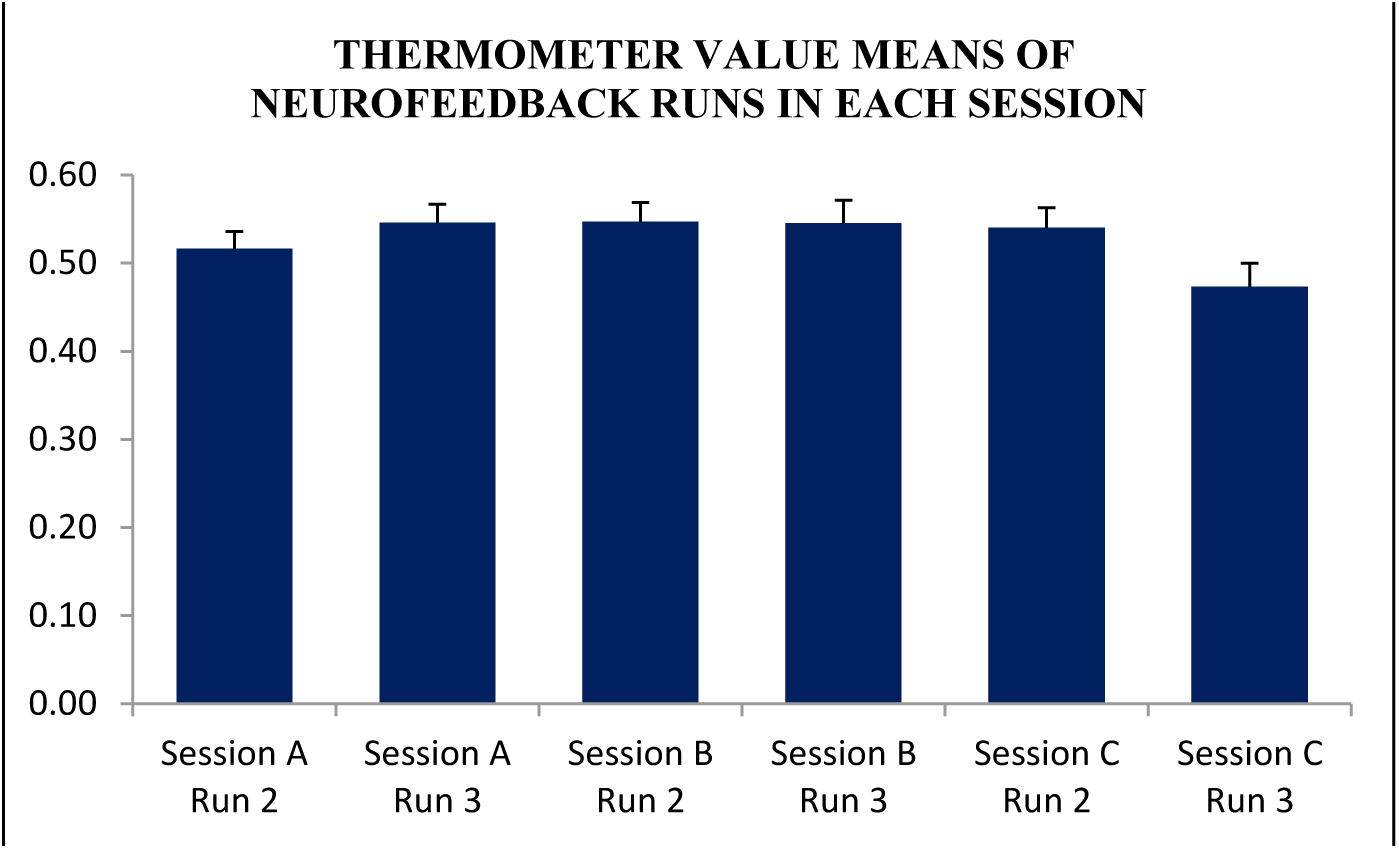
Neurofeedback Success: Thermometer Scale Position. NF participants were successful in controlling the neurofeedback thermometer on average with positive feedback around the 50% mark of the thermometer scale (0-100% range). Participants’ neurofeedback success occurred already in the first session and was stable throughout further sessions with a slight drop in the final active run. FRIEND’s moving target algorithm means that after successfully reducing rSATL-posterior SC connectivity, it could be increasingly difficult to further successfully reduce it in subsequent training runs.

**Supplementary Figure 5.**
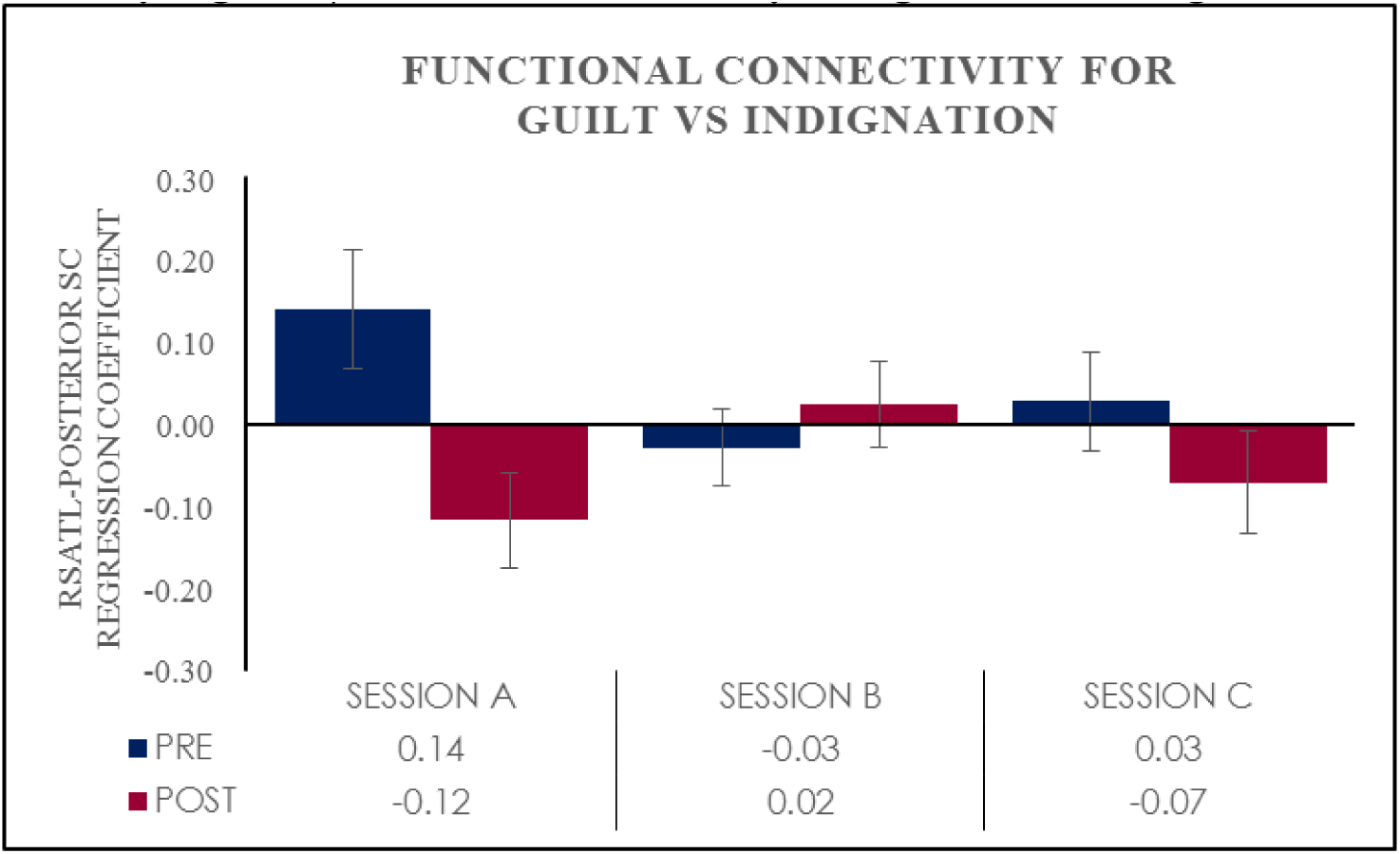
Functional connectivity changes over training sessions. Change in functional connectivity between rSATL and posterior SC in the guilt vs indignation condition, measured as Cohen’s D for regression coefficient means with standard errors for time series pre- and post-rtfMRI neurofeedback training in n=18 participants plotted for each neurofeedback session.

